# Local prevalence of transmissible SARS-CoV-2 infection: an integrative causal model for debiasing fine-scale targeted testing data

**DOI:** 10.1101/2021.05.17.21256818

**Authors:** George Nicholson, Brieuc Lehmann, Tullia Padellini, Koen B Pouwels, Radka Jersakova, James Lomax, Ruairidh E King, Ann-Marie Mallon, Peter J Diggle, Sylvia Richardson, Marta Blangiardo, Chris Holmes

## Abstract

Targeted surveillance testing schemes for SARS-CoV-2 focus on certain subsets of the population, such as individuals experiencing one or more of a prescribed list of symptoms. These schemes have routinely been used to monitor the spread of SARS-CoV-2 in countries across the world. The number of positive tests in a given region can provide local insights into important epidemiological parameters, such as prevalence and effective reproduction number. Moreover, targeted testing data has been used inform the deployment of localised non-pharmaceutical interventions. However, surveillance schemes typically suffer from ascertainment bias; the individuals who are tested are not necessarily representative of the wider population of interest. Here, we show that data from randomised testing schemes, such as the REACT study in the UK, can be used to debias fine-scale targeted testing data in order to provide accurate localised estimates of the number of infectious individuals. We develop a novel, integrative causal framework that explicitly models the process underlying the selection of individuals for targeted testing. The output from our model can readily be incorporated into longitudinal analyses to provide local estimates of the reproduction number. We apply our model to characterise the size of the infectious population in England between June 2020 and January 2021. Our local estimates of the effective reproduction number are predictive of future changes in positive case numbers. We also capture local increases in both prevalence and effective reproductive number in the South East from November 2020 to December 2020, reflecting the spread of the Kent variant. Our results illustrate the complementary roles of randomised and targeted testing schemes. Preparations for future epidemics should ensure the rapid deployment of both types of schemes to accurately monitor the spread of emerging and ongoing infectious diseases.

## Introduction

The spread of the new severe acute respiratory syndrome coronavirus 2 (SARS-CoV-2) and the ensuing outbreaks of coronavirus disease 2019 (COVID-19) have placed a significant burden on public health in the United Kingdom (UK). As of 12 April 2021, the number of people who have died within 28 days of a positive SARS-CoV-2 test was 127,100 [1, 2]. In response to the ongoing epidemic, the UK government has implemented a number of non-pharmaceutical interventions (NPIs) to reduce transmission of SARS-CoV-2, ranging from localised measures, such as the closures of bars and restaurants, to full national lockdowns [3]. The localised measures have been employed through a regional tier system, with areas being placed under varying levels of restrictions according to data such as the number of positive polymerase chain reaction (PCR) tests returned there over a seven-day interval (or local *weekly* positive tests) [4]. Following a third national lock-down that began on the 6th January 2021, the UK is currently undergoing a staged relaxation of restrictions [5]. Accurate local measures of prevalence and incidence are needed to assess the need for any changes to this plan and importantly to measure the relative impact of the individual stages thereby providing crucial information for future waves and pandemics both in the UK and more globally.

In the UK, there are two major, ongoing studies that undertake randomised testing to provide an insight into the prevalence of SARS-CoV-2. Since April 2020, the Office for National Statistics (ONS) COVID-19 Infection Survey (CIS) tests a random sample of people living in the community with longitudinal follow-up. [6]. The survey is designed to be representative of the UK population, with individuals aged 2 years and over in private households randomly selected from address lists and previous ONS surveys, though it does not explicitly cover care homes, the sheltering population, student halls or individuals currently being hospitalised. The REal-time Assessment of Community Transmission (REACT) study is a second nationally representative prevalence survey of SARS-CoV-2 based on repeated cross-sectional samples from a representative subpopulation defined via (stratified) random sampling from England’s National Health Service patient register [7]. Importantly, both surveys recruit participants regardless of symptom status and are thus able to largely avoid issues arising from ascertainment bias when estimating prevalence. The ONS CIS uses multilevel regression and poststratification to account for any residual ascertainment effects due to non-reponse [6] while REACT uses survey weights for this purpose.

While randomised surveillance testing readily provides an accurate statistical estimate of prevalence of PCR positivity, precision can be low at finer spatiotemporal scales (e.g. the lower tier local authority (LTLA) level), even in large studies such as the ONS CIS and REACT surveys. The major goal there is to unlock the information in non-randomised testing under arbitrary, unknown ascertainment bias. While we expect the methods to apply broadly, here we focus in on Pillar 1 and Pillar 2 PCR tests conducted in England between 31st May 2020 and 24th January 2021 (lateral flow device, LFD, tests are not included; further details in Methods–*Data*). As Pillar 1 tests refer to *“all swab tests performed in Public Health England (* PHE*) labs and National Health Service (* NHS*) hospitals for those with a clinical need, and health and care workers”*, and Pillar 2 comprises *“swab testing for the wider population”*, Pillar 1+2 testing has more capacity than the randomised programs, but the protocol incurs ascertainment bias as those at elevated risk of being infected are tested, such as frontline workers, contacts traced to a COVID-19 case, or the sub population presenting with COVID-19 symptoms, such as loss of taste and smell [8]. Hence, raw prevalence estimates from Pillar 1+2 data (as a proportion of tested population) will tend to be biased upwards and cannot directly be used to estimate the unknown infection rate in a region (in contrast, as a proportion of the whole population the bias is downwards as not all infected individuals in the area are captured). Also they tend not to capture asymptomatic infection, while there is evidence that asymptomatic individuals can contribute to spread of the virus [9, 10].

Combining data from multiple surveillance schemes can improve estimates for prevalence. For example, Manzi et al. incorporate information from multiple, biased, commercial surveys to provide more accurate and precise estimates of smoking prevalence in local authorities across the east of England [11]. A number of geostatistical frameworks for infectious disease modelling based on multiple diagnostic tests have been developed [12, 13, 14]. These accommodate different sources of heterogeneity among the tests to deliver more reliable and precise inferences on disease prevalence.

To understand the ascertainment bias problem and a statistical approach to correction, it is helpful to consider a simplified causal model for Pillar 1+2 data. This is represented by a directed acyclic graph (DAG), shown in Figure 1(a), that charts the dependencies of an individual from infection status to test result. The circles indicate the binary (Yes/No) states of an individual. The DAG characterises the joint distribution of the major factors leading to the observed data. Throughout the paper we use the term *targeted* testing data to refer to data gathered under some ascertainment process distinct from (stratified) random sampling, with an exemplar being selection for testing of the subpopulation with COVID-19 symptoms, which comprises a sizeable proportion of Pillar 1+2 tests. The Pillar 1+2 DAG can be compared to that of a randomised surveillance study (shown in Figure 1(b)). The randomised nature of the test allocations in REACT renders Tested conditionally independent of Symptoms given Infected, yielding unbiased estimates of Infection rates. The DAG explicitly characterises statistically why we cannot use Pillar 1+2 data directly. The DAG also points to a potential solution if the statistical dependencies as indicated by the arrows in Figure 1(a) can be modelled, then Pillar 1+2 data *can* be used. In this paper, we describe an approach allowing characterisation and adjustment for the ascertainment bias inherent in Pillar 1+2 data.

**Figure 1:**
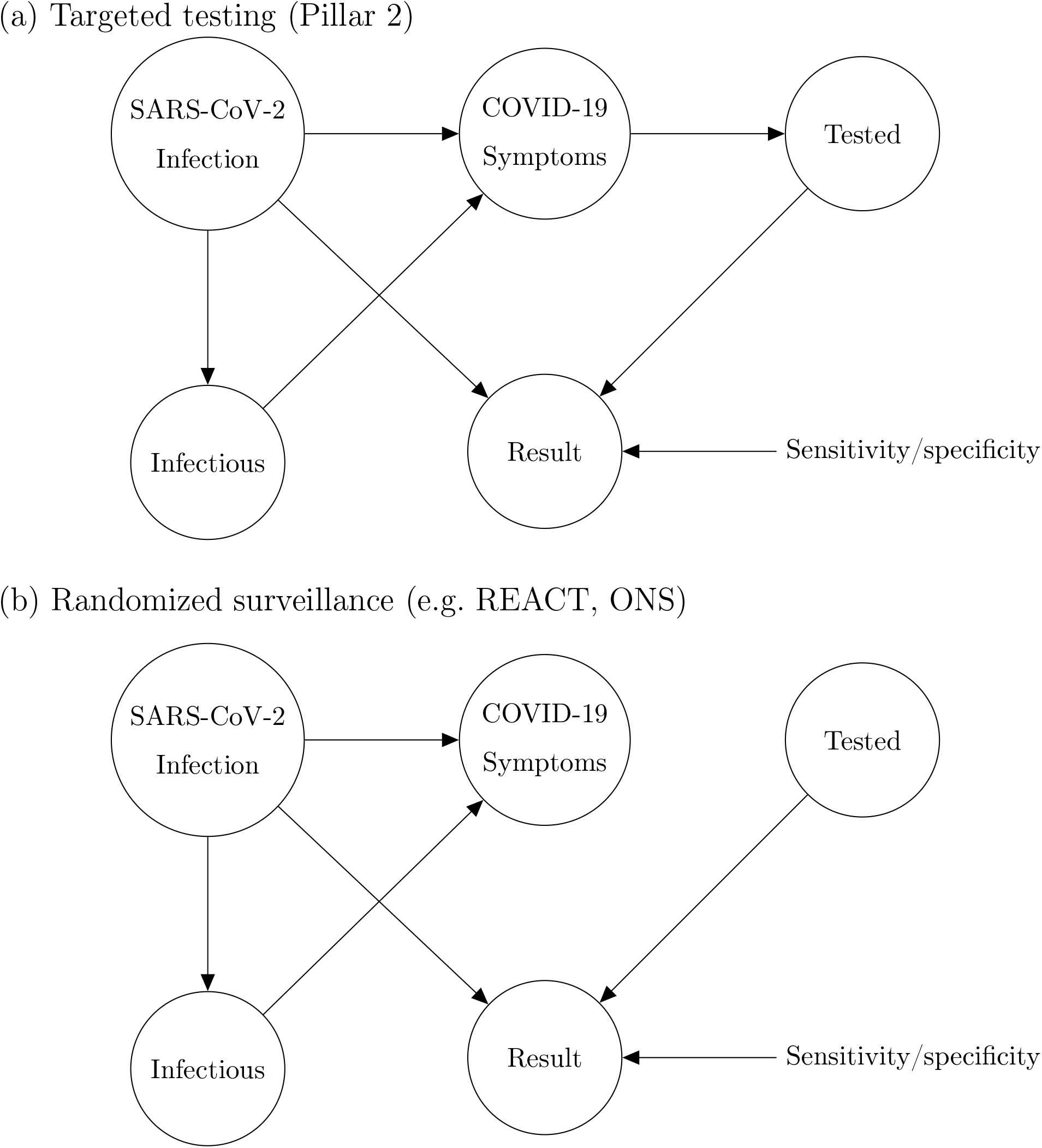
DAGs representing causal models underlying SARS-CoV-2 swab testing data for (a) Targeted test-and-trace data (Pillar 1+2); and (b) Randomised surveillance data (e.g. REACT). In (b), randomisation breaks the causal link between COVID-19 symptoms and swab testing. The nodes represent binary (yes/no) states for an individual in the relevant population.

In addition to prevalence, there are a number of epidemiological parameters that may be useful for informing localised NPIs. For example, one particular variable of interest is the (time-varying) effective reproductive number *ℛ*_*t*_, defined roughly as the average number of infections caused by an infectious individual; when *ℛ*_*t*_ *>* 1, the epidemic will continue to spread. Estimation of these parameters relies on careful mathematical modelling supported by relevant data such as surveillance testing results or number of hospitalisations [15]. Incorporating multiple sources of data can produce more reliable parameter estimates [16], though doing so in a computationally efficient manner may be nontrivial [17]. For example, a stochastic epidemic model of the 2009 influenza outbreak in Finland that used data on hospitalisations, lab tests and vaccination data provided insights into the time-varying nature of *ℛ*_*t*_ but took over a month of compute time to run [18]. Given the time-sensitive nature of the current epidemic, one important modelling consideration is the timely inference of parameters; the work we present here has been developed with both accuracy and computational efficiency in mind.

The current pandemic has spurred the development of a number of models that also aim to incorporate multiple sources of data in order to estimate important epidemiological parameters, in particular *ℛ*_*t*_ [19]. Abbott et al. [20] generate daily estimates of *ℛ*_*t*_ at a national and PHE region level by incorporating case counts and death notifications, building on a model to estimate *ℛ*_*t*_ from incidence time series [21]. Birrell et al. [22] estimate daily, PHE regional *ℛ*_*t*_ using ONS CIS data, death notifications and serological data within an age-stratified transmission model. Colman et al. [23] develop methods to combine Pillar 1+2 alongside ONS CIS data to estimate the proportion of infections that result in a positive diagnosis, outputting estimates of the true incidence of infections over time. Methods have also been proposed to use serological data to adjust for the effects of biases in testing data in order to estimate the infection to fatality ratio [24, 25].

The research referenced so far infers epidemiological parameters at spatially coarse scales, such as PHE region. To extend this body of work, we focus here on accurate estimation of prevalence (along with other epidemiological parameters) at a more local level, such as the LTLA.

There are two very useful websites providing LTLA-level up-to-date estimates and predictions of some epidemiological parameters (but not prevalence).^1^ A team at Oxford has produced a local Covid map^2^ as part of the Royal Society Data Evaluation and Learning for Viral Epidemics (DELVE) initiative.^3^ Their methods take as input Pillar 1+2 daily counts and commuter flow data, and output local estimates and predictions of *ℛ*_*t*_ and positive case numbers. An Imperial College team has produced a COVID-19 UK map and table.^4^ Their methodology takes as input: daily cases data, weekly deaths data, as well as daily infections from the ONS CIS and REACT data sets [26]. They output estimates and predictions of *ℛ*_*t*_, positive case numbers, and change in new infections. Their results are based on the `epidemia` software [27], which is an extension of the Bayesian semi-mechanistic model introduced in [28], though detailed methods are not yet available. We have downloaded their *ℛ*_*t*_ estimates and find a high level of consistency in local *ℛ*_*t*_ estimates between our model and theirs. A group from Lancaster University has estimated daily case prevalence (proportion of infected population), incidence, and *ℛ*_*t*_ at the Local Authority District level in England (315 areas in total) by building an epidemic model incorporating measures of human mobility and Pillar 1 and 2 tests across England [29]. An important aspect of this approach is that it assumes each infection is eventually reflected in the Pillar 1 and 2 case reports and so does not account for possible ascertainment bias in targeted testing. Furthermore, the model requires substantial computational resources to obtain timely estimates.

Within this urgent and fast developing area of research, it is clearly important to define the aspects in which our method contributes novelty. Firstly, we have developed methods to infer local prevalence, *I*_*t*_, accurately from targeted testing data. Here we work with weekly period prevalence, and explicitly target the number of infectious individuals via a correction to the estimated PCR-positive numbers. This is all novel and important in its own right – being able to estimate local prevalence accurately from targeted testing data adds an important facet to existing COVID-19 monitoring capabilities. Second, our method outputs bias-adjusted cross-sectional prevalence likelihoods *p*(*n*_*t*_ of *N*_*t*_ | *I*_*t*_), where *n*_*t*_ and *N*_*t*_ are positive and total targeted test counts. This allows prevalence information from targeted data to be coherently embedded in a modular way into complex spatiotemporal epidemiological models, including those synthesising multiple data types. We exemplify this by implementing an Susceptible-Infectious-Recovered (SIR) model around our ascertainment model likelihood. Third, our local ascertainment model is based on targeted testing data alone with, uniquely to our knowledge, both the number of positive *and total* tests being modelled (*n*_*t*_ *and N*_*t*_). This has two important benefits: spatiotemporal variation in testing uptake and capacity is explicitly conditioned on (via *N*_*t*_), and differential test specificity and sensitivity can be be naturally incorporated into our causal ascertainment model.

## Results

### Correcting for ascertainment bias in targeted testing data

Figure 2(a-b) displays the percentage of positive Pillar 1+2 tests (as a proportion of those tested) against accurate prevalence estimates from the REACT study, showing a clear upward bias (each point corresponds to a single LTLA). Here we introduce a bias-correction method that aims to provide accurate estimates of prevalence at the local level as displayed in Figure 2(c-d), based on the posterior cross-sectional prevalence *p*(*I*_*t*_ | *n*_*t*_ of *N*_*t*_).

**Figure 2:**
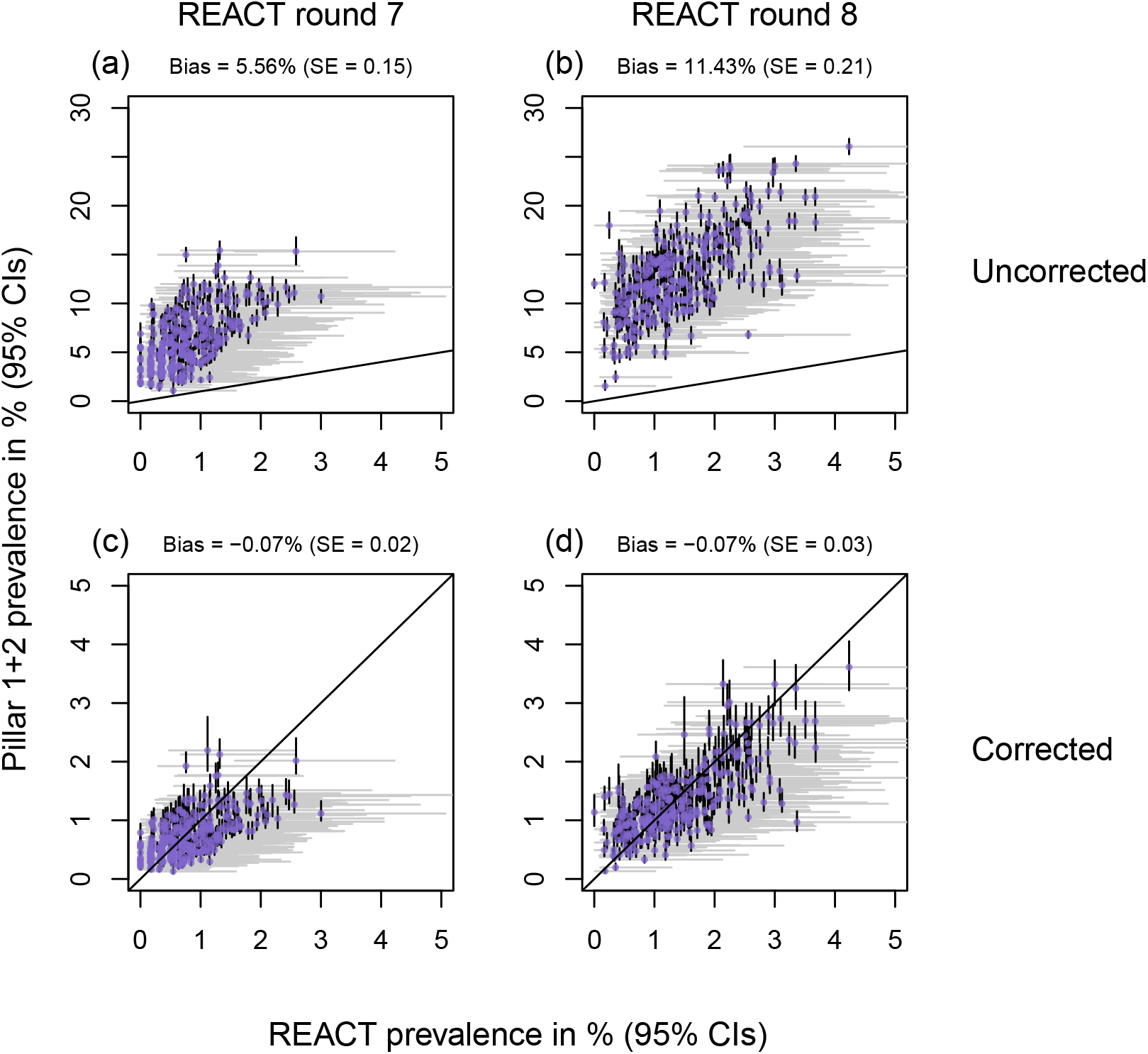
Uncorrected and corrected Pillar 1+2 PCR-positive prevalence estimates against (gold-standard) REACT estimates from randomised surveillance. Each point corresponds to an LTLA. Each scatter plot compares Pillar 1+2 prevalence estimates against unbiased estimates from the REACT study. Panels (a,c) show REACT round 7 data (13th Nov - 3rd Dec 2021), and (b,d) show round 8 (6th-22nd Jan 2021). Uncorrected results are shown in panels (a-b) and bias-corrected cross-sectional estimates in (c-d). Horizontal grey lines are 95% exact binomial confidence intervals from the REACT data. Vertical black lines in panels (a) and (b) are 95% exact binomial confidence intervals for from the raw, non-debiased Pillar 1+2 data. Vertical black lines in panels (c) and (d) are 95% posterior credible intervals from the debiased Pillar 1+2 data. Neither set of prevalence estimates has been corrected for false positives/negatives. Note that in panels (c) and (d), the CI widths are systematically tighter for the debiased Pillar 1+2 compared to the REACT data, pointing to the useful information content in debiased Pillar 1+2 data.

With reference to the causal DAG in figure 1, we define the essential bias parameter, *δ*, as

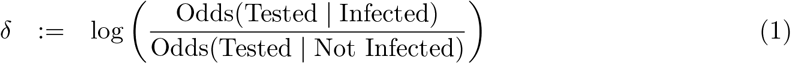

i.e. the log odds ratio of being tested in the infected versus non-infected populations. Larger values of *δ* generally correspond to higher levels of ascertainment bias, i.e. a higher chance of an infected individual being selected for testing, relative to a non-infected.

Our approach combines randomised surveillance data (REACT) and targeted surveillance data (Pillars 1 and 2) to infer *δ* at the coarse geographical level (PHE region). We then integrate this information by specifying a temporally smooth empirical Bayes (EB) prior on *δ*_1:*T*_, applied to each constituent local region (LTLA) in the local prevalence analyses. Figure 3 shows the resulting EB priors on *δ*; there is potentially more variation in *δ* across regions early on in the sampling period (pre-September 2020), though the prior credible intervals (CIs) are quite broad and often overlapping. The data provide more information on *δ* from October 2020 onwards, and there is a consistent upward trend for all nine PHE regions.

**Figure 3:**
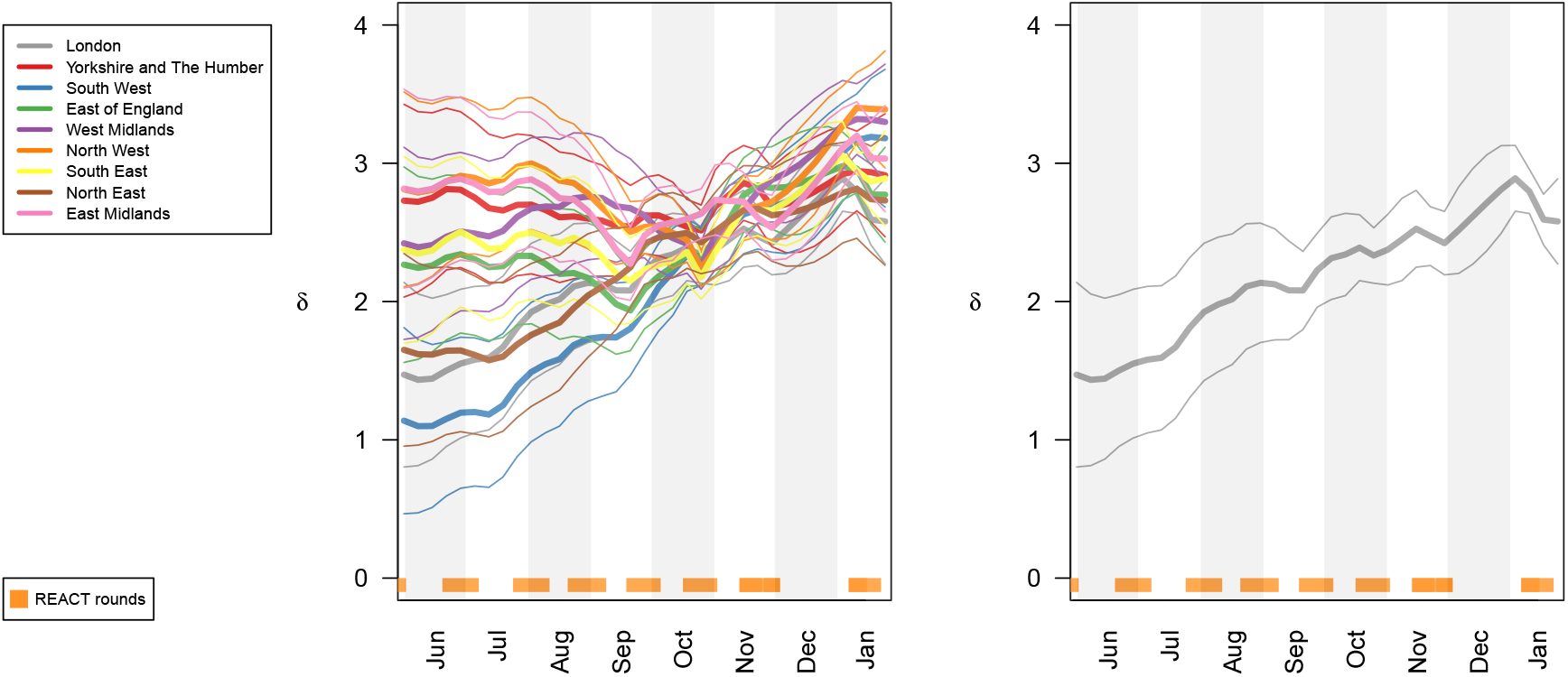
Smooth EB priors on bias parameters *δ*_1:*T*_. Left: Showing heterogeneous bias across the nine PHE regions. Right: London only. 95% CIs shown. Note that *δ* is the log odds ratio, so for example *δ* = 3 implies that the odds of being Tested are *e*^3^ *≈* 20 times higher in Infected compared to Not Infected individuals

### Cross-sectional local prevalence from targeted testing data

#### De-biased likelihood for modular sharing of prevalence information

Equipped with a coarse-scale (PHE-region level) EB prior on bias *δ* we evaluate a fine-scale (LTLA-level) *δ*-marginalized likelihood of the form *p*(*n*_*t*_ of *N*_*t*_ | *I*_*t*_, 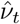), as described at (17) in Methods–*Cross-sectional inference on local prevalence*. This de-biased prevalence likelihood can be readily exported and modularly incorporated into more complex models, as we illustrate below in Results–*Longitudinal local prevalence and transmission*.

#### Cross-sectional prevalence posterior

The *δ*-marginalized likelihood can be inputted directly into cross-sectional Bayesian inference, outputting the prevalence posterior *p*(*I*_*t*_ | *n*_*t*_ of *N*_*t*_, 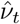) for each time point at which such count data are available. Figure 4 plots these cross-sectional prevalence posteriors beneath the raw counts for a subset of LTLAs across the nine PHE regions. REACT sampling periods are plotted at the base of each panel, and local prevalence estimates from REACT round 7 (November 2020) and round 8 (January 2021) are also superimposed. The corrected cross-sectional prevalence estimates are consistent with the gold-standard REACT estimates, but are more precise, as expected from Bayesian principles of data synthesis. Inspecting the bias-corrected estimates within LTLA across time points, they tend to show relatively wider 95% CIs in time intervals between REACT sampling rounds (particularly in the December 2020 period between round 7 and 8) reflecting the dependence on the REACT data for good inference.

**Figure 4:**
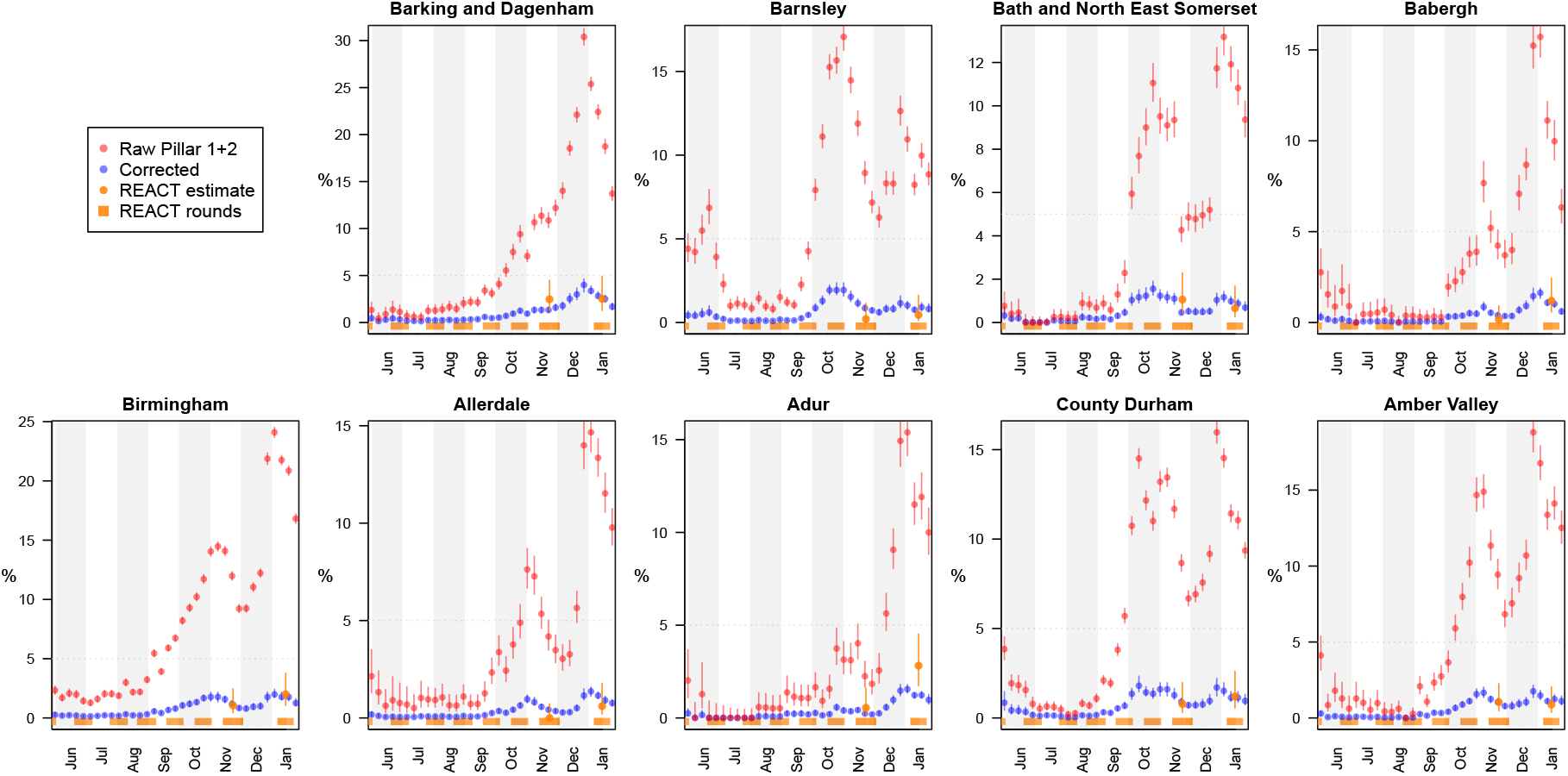
LTLA-level prevalence estimates: Raw Pillar 1+2 estimates, cross-sectionally corrected Pillar 1+2, and gold-standard REACT estimates (see legend). For each of the nine PHE regions, we present the constituent LTLA whose name is ranked top alphabetically.

### Longitudinal local prevalence and transmission

The cross-sectional de-biased likelihood can be introduced modularly into a wide variety of down-stream epidemiological models. We illustrate this by using the likelihood as an input to a simple SIR epidemic model (see Methods–*Full Bayesian inference under a stochastic* SIR *epidemic model* and Figure 12). Figure 5(a) plots estimated prevalence against *ℛ*_*t*_ number at the most recent time point (the week of 2021-01-24), with each point corresponding to a single LTLA. The scatter plot provides a quick visual representation of regions where transmission rates and/or prevalence are relatively high – to illustrate, we label five LTLAs with high prevalence and/or *ℛ*_*t*_ estimates. The estimated longitudinal prevalence and *ℛ*_*t*_ for this subset of LTLAs (Figure 5(b-c)) can help further to characterise the longitudinal dynamics of prevalence and transmission in the time interval leading up to 2021-01-24, in particular showing the estimated rate of change in prevalence and separately indicating whether *ℛ*_*t*_ is increasing or decreasing.

**Figure 5:**
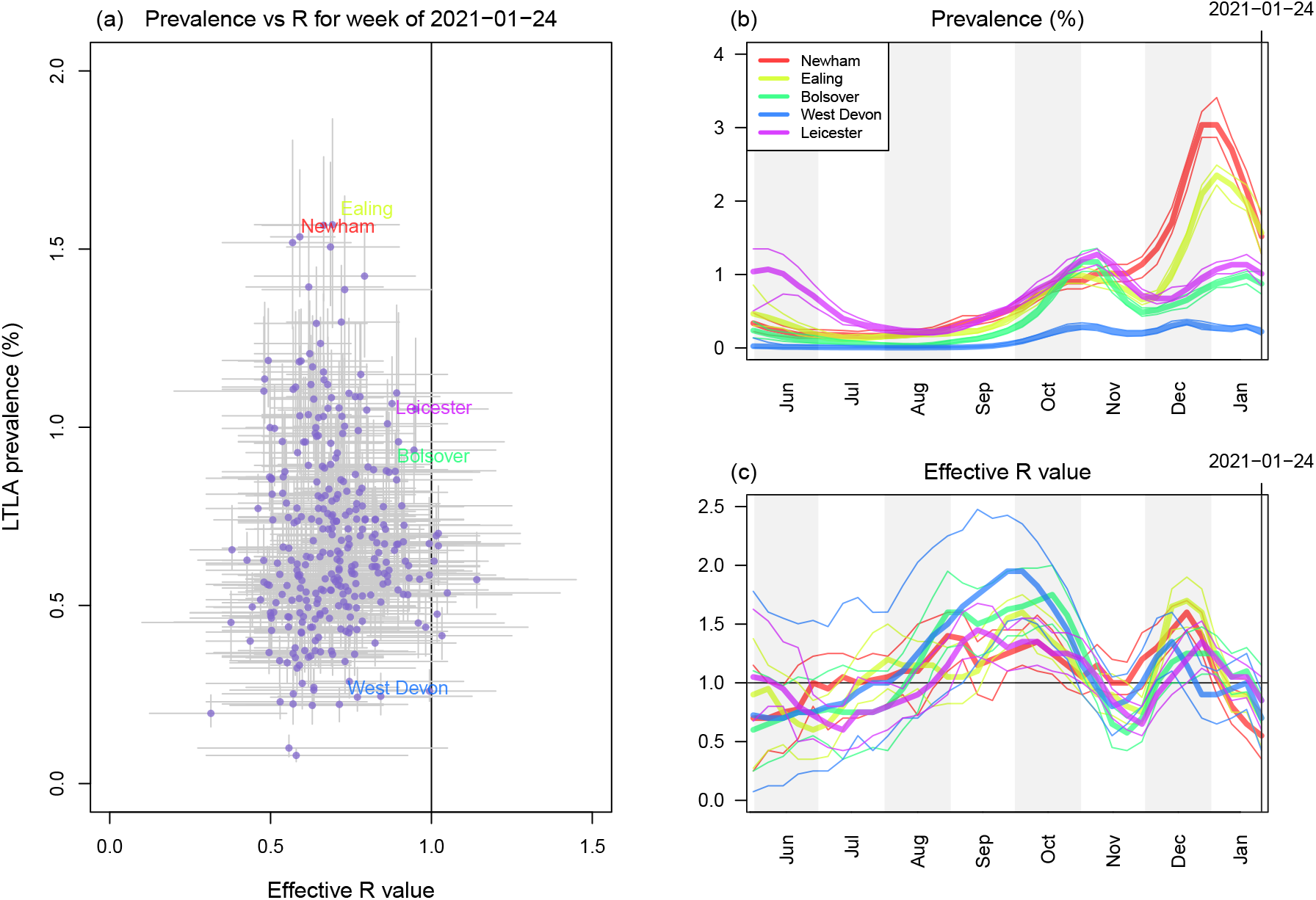
Outputs of longitudinal local prevalence model. (a) Scatterplot of prevalence against effective R number (each point corresponds to one LTLA). (b) Longitudinal posteriors for prevalence at a selection of LTLAs. (c) Longitudinal posteriors for *ℛ*_*t*_ at a selection of LTLAs.

Figures 6 and 7 display spatiotemporal local prevalence and *ℛ*_*t*_ respectively, using a sequence of weekly maps, with each LTLA coloured according to its weekly prevalence estimate. Zoom-in boxes display the local fine-scale structure for expanded areas including London and the North West.

**Figure 6:**
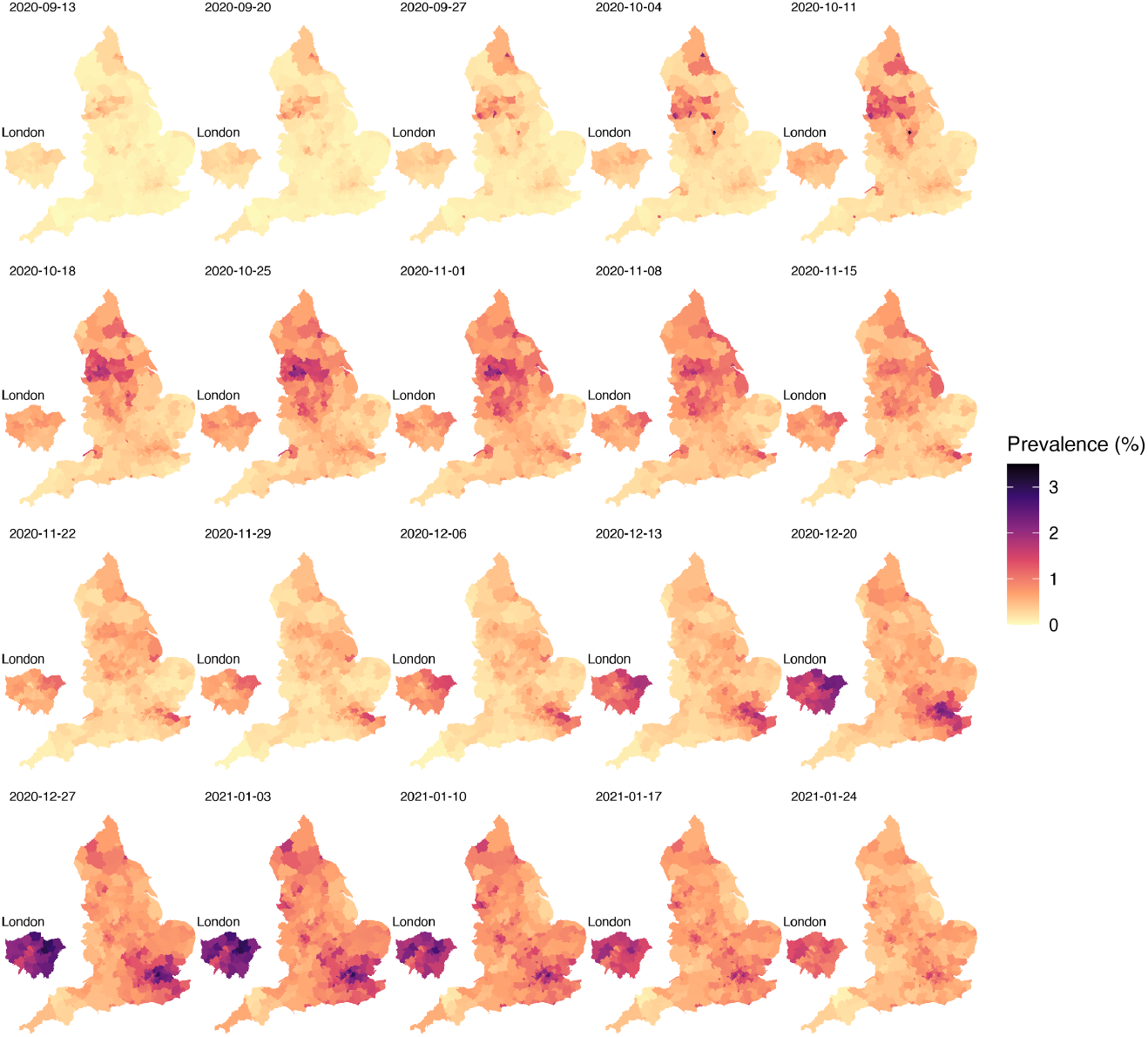
Longitudinal maps of estimated local prevalence from 13th September 2020 to 24th January 2021.

**Figure 7:**
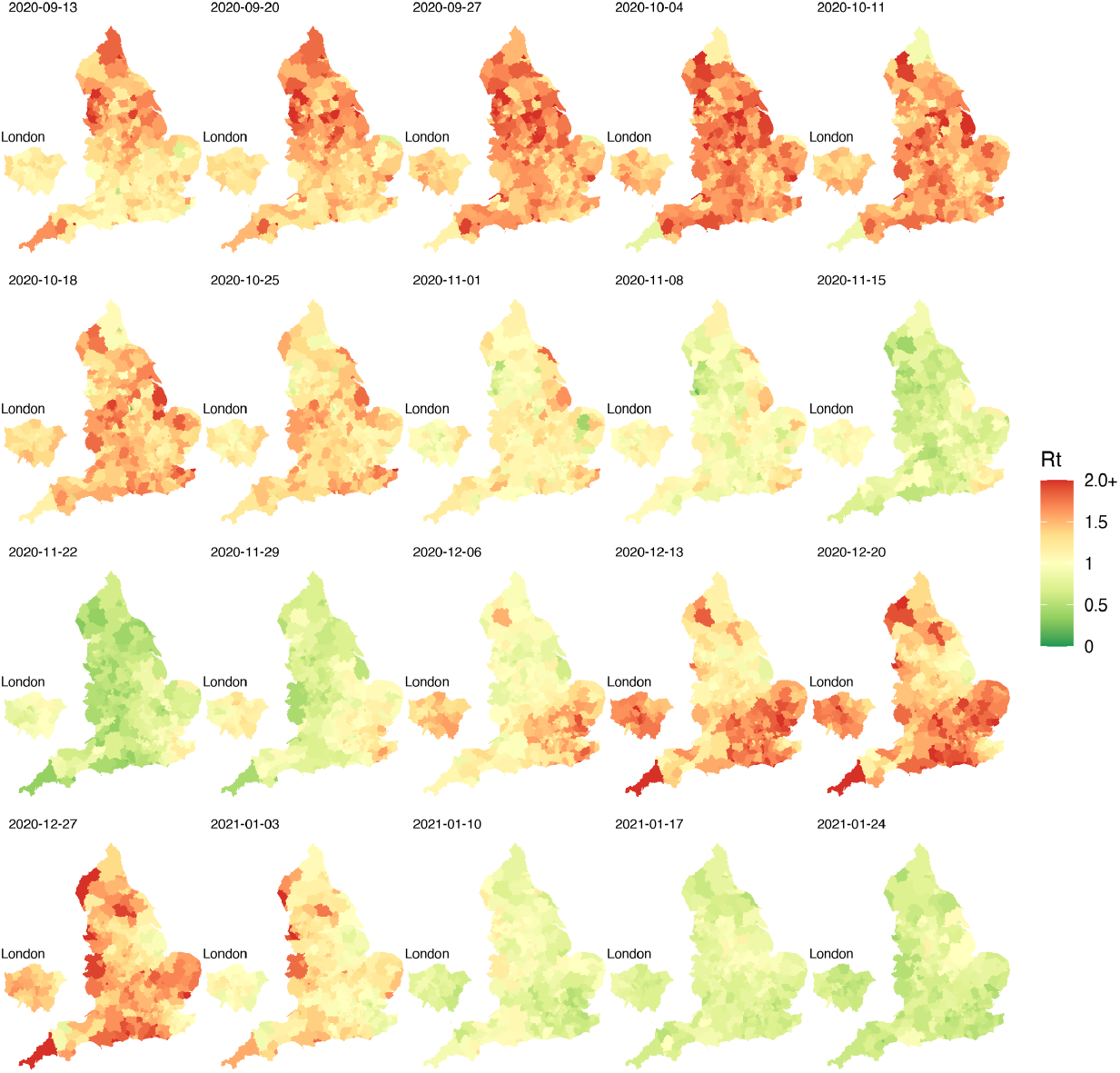
Longitudinal maps of estimated local *R*_*t*_ from 13th September 2020 to 24th January 2021.

### Relating local prevalence and transmission to spread of the UK variant

One striking feature of the maps in Figure 6 is the increasing prevalence in the London area throughout November to December 2020. This is consistent with the known arrival of the UK variant of concern (VoC) 202012/01 (lineage B.1.1.7), that emerged in the South East of England in November 2020 and which has been estimated to have a 43–90% higher reproduction number than preexisting variants [30].

We investigate this hypothesis similarly to [30], by characterising the relationship between estimated local *ℛ*_*t*_ and the frequency of VoC 202012/01, as approximated by the frequency of S gene target failure (SGTF) in the Taqpath sequencing assay used over this time period [31]. Figure 8 illustrates the spatial distributions of VoC 202012/01 against estimated prevalence and estimated *ℛ*_*t*_ from mid-November 2020 to mid-December 2020. The increase in frequency of the VoC was initially isolated to the South-East but then spread outwards, accompanied by a corresponding increase in both local estimated prevalence and *ℛ*_*t*_. We observe a strong positive association between the local VoC frequency and estimated local *ℛ*_*t*_, consistent with the increased transmissibility identified in [30].

**Figure 8:**
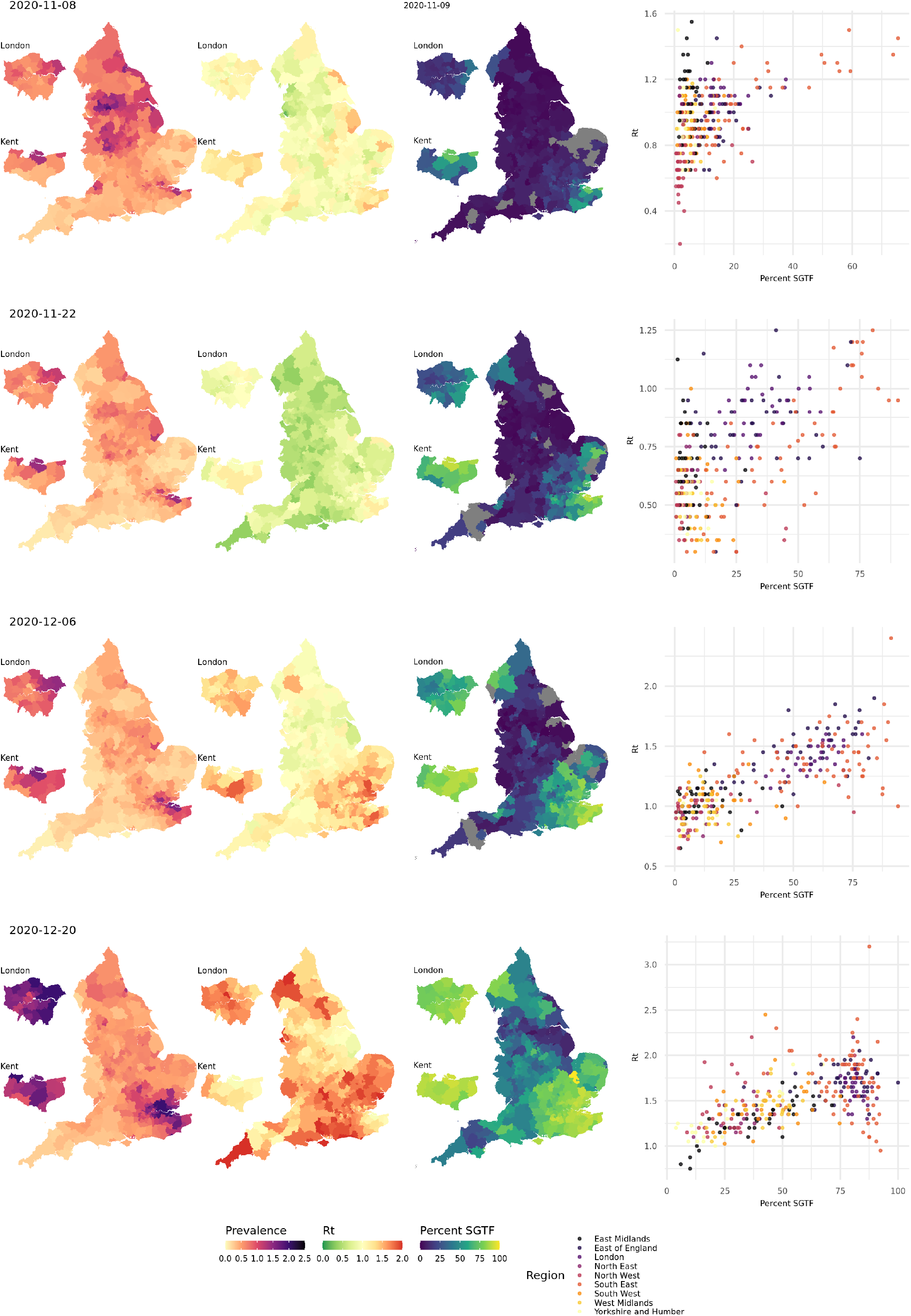
Maps of estimated local prevalence (left), estimated local *R*_*t*_ (middle), and frequency of S gene target failure (SGTF; right), and scatter plot of SGTF frequency against estimated *R*_*t*_.

### Validation 1 (accuracy)

We qualitatively assess the performance of de-biased fine-scale (LTLA-level) prevalence estimates by measuring how well they predict LTLA-level REACT data. The validation is best described in terms of coarse-scale REACT training data and contemporaneous fine-scale REACT test data. The training data inputted are REACT PHE region-level and Pillar 1+2 LTLA-level positive (and number of) test counts for the week at the centre of the corresponding REACT round to be predicted. The test data are REACT LTLA-level positive (and number of) test counts aggregated across the relevant REACT sampling round. Figure 2(c-d) visually compares cross-sectional LTLA prevalence estimates from de-biased targeted data (i.e. based only on the training data) with accurate gold-standard estimates from REACT LTLA-level test data. The average estimated bias is reduced to low levels for comparisons with both REACT round 7 (−0.07%, SE = 0.02) and round 8 (−0.07%, SE = 0.03).

### Validation 2 (prediction)

The effective reproduction number, *ℛ*_*t*_, measures whether the number of infectious individuals is increasing, *ℛ*_*t*_ *>* 1, or decreasing, *ℛ*_*t*_ *<* 1, in the population at time point *t*. Figure 9 compares LTLA *ℛ*_*t*_ estimates with the future change in local case numbers. For validation purposes, here we are doing one-step-ahead at a time prediction and comparing predictions with out-of-training-sample observed statistics (fold change in raw case numbers from baseline). The results are stratified according to baseline case numbers, and we examine predictions one week and two weeks ahead. Each point corresponds to an (LTLA, week) pair, and the results are for the period 2020-10-18 - 2021-01-24. Across each of the six scenarios presented, there is strong evidence of association between *ℛ*_*t*_ and future change in case numbers (*p <* 2 *×* 10^−14^). The strength of association between *ℛ*_*t*_ and one week ahead case numbers has Spearman’s *ρ* = 0.68 for the high baseline case group (>500 cases per 100,000), decreasing to *ρ* = 0.31 in the low baseline group (≤ 200 cases per 100,000). The association remains strong when predicting caseloads two weeks ahead, with for example *ρ* = 0.69 (Spearman) for the high baseline case group.

**Figure 9:**
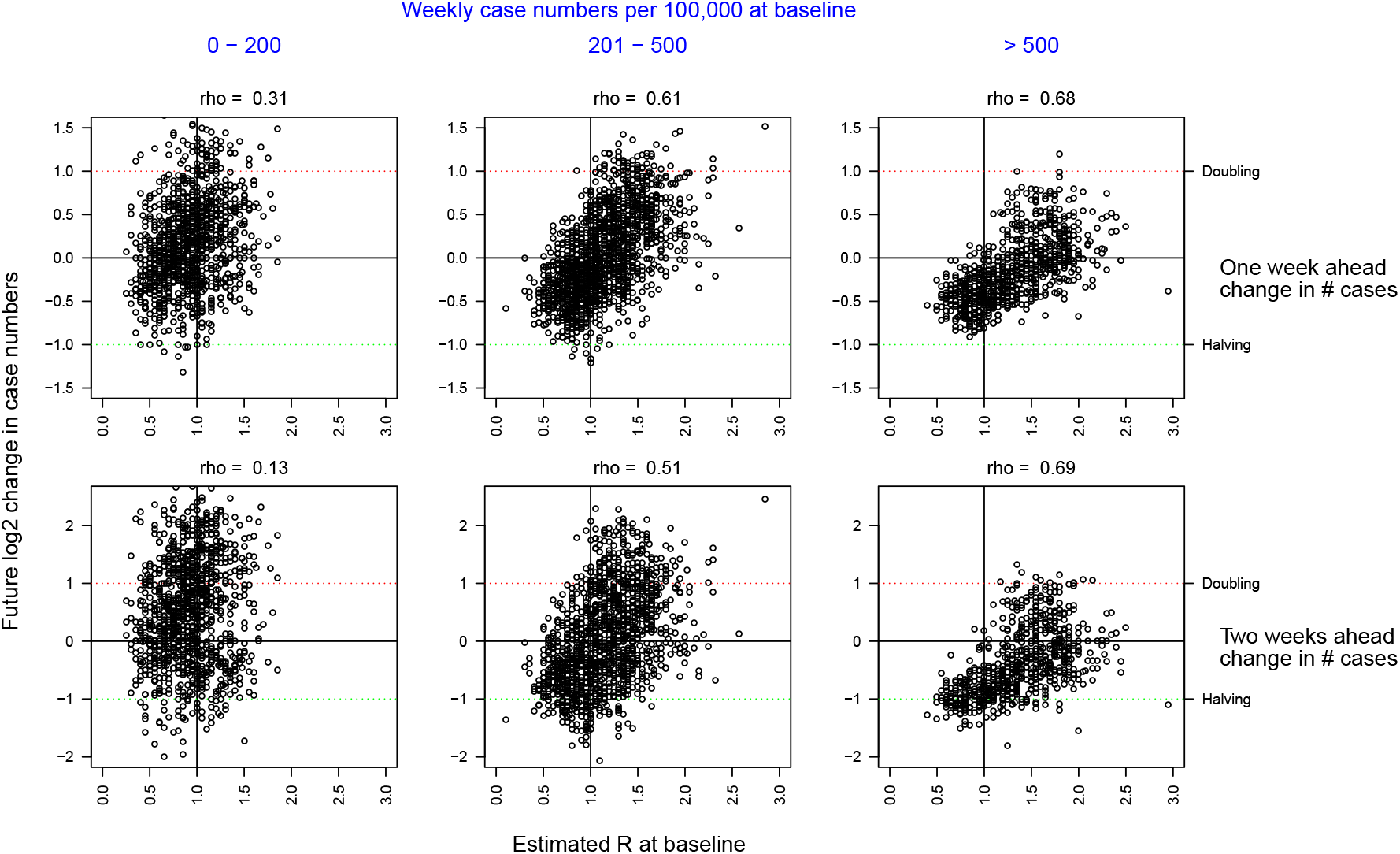
Predicting future change in case numbers from current estimated *ℛ*_*t*_. Each point corresponds to an (LTLA, week) pair, predicting future case numbers in the LTLA using *ℛ*_*t*_ for that week. Future case numbers are represented by forward-in-time log_2_ fold change log_2_(*n*_*t*+*k*_*/n*_*t*_). Case data underlying the plot are from the period 2020-10-18 - 2021-01-24. Note the number of points in each column differs based on how many LTLA-week pairs have baseline case numbers in the intervals in blue shown at the top of the plot.

### Validation 3 (external)

We extracted estimates of effective reproduction number *ℛ*_*t*_ based on our de-biasing model likelihood implemented within a standard SIR model, illustrated in Figure 12. We compare the results to the local *ℛ*_*t*_ estimates outputted by at the Imperial College COVID-19 website.^5^ A cross-method comparison of longitudinal traces of *ℛ*_*t*_ for a subset of LTLAs is shown in Figure 10. Encouragingly for both approaches, the estimates generally display good concordance, with credi-ble intervals overlapping appropriately, despite being based on different data and models.^6^.

**Figure 10:**
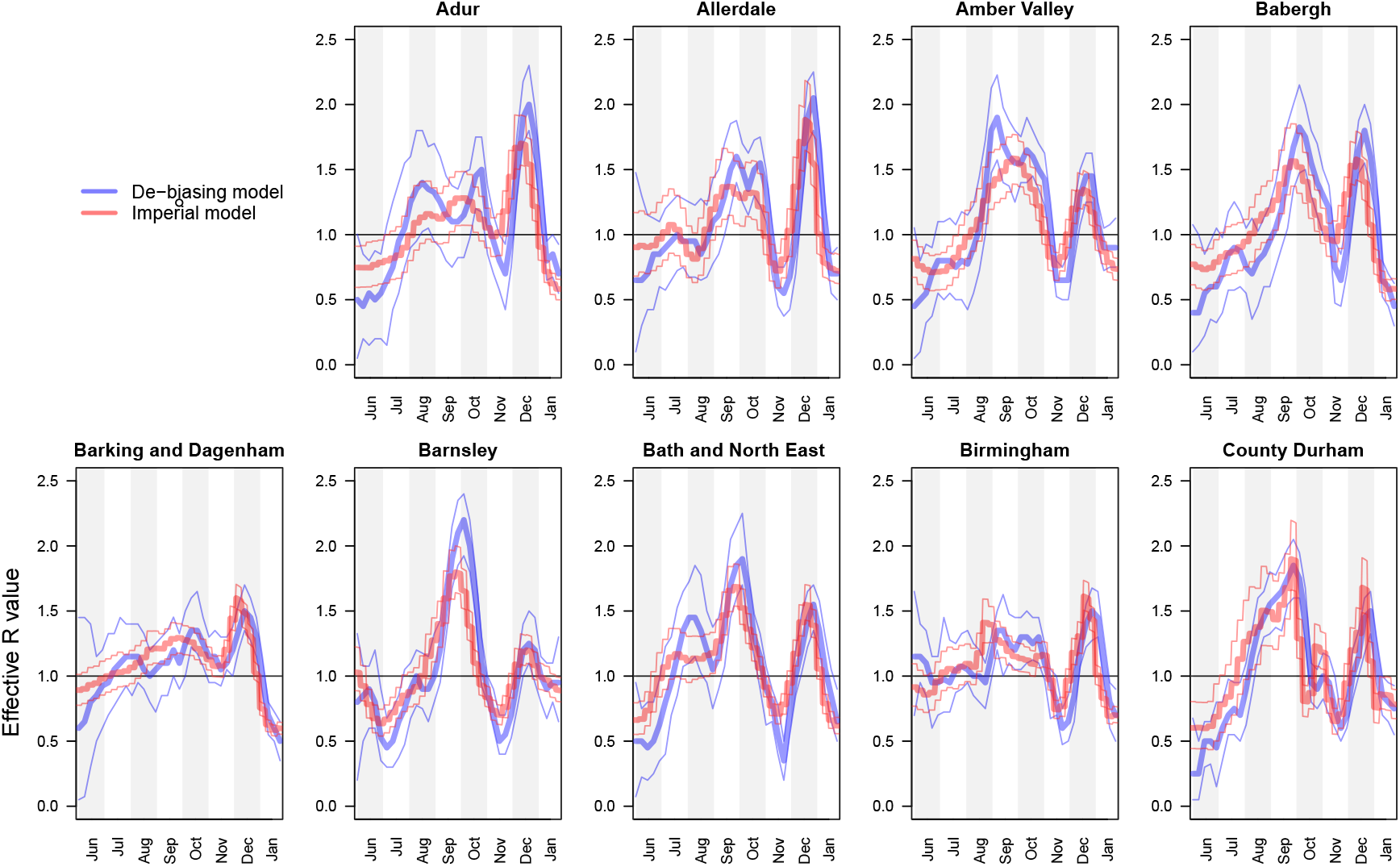
Comparison of *ℛ*_*t*_ estimates between de-biasing model and Imperial model [32]. For each of the nine PHE regions, we present the constituent LTLA whose name is ranked top alphabetically.

## Methods

### Observational models for surveillance data

The primary target of inference is prevalence, *I* out of *M*, being the unknown number of individuals infected at a particular time-point in the local population of known size *M*. Our method estimates two types of prevalence: 1) the number of individuals that would test PCR positive (*Ĩ*), and the number of individuals that are infectious (*I*); see Methods–*Focusing prevalence on the infectious subpopulation*. We clarify below the distinction between the PCR positive and infectious subpopulations, and how we target the latter.

#### Randomised surveillance data, *u* of *U*

Suppose that out of a total *U* randomised surveillance (e.g. REACT, ONS CIS) tests, we observe *u* positive tests. The randomised testing (e.g. REACT, ONS CIS) likelihood is

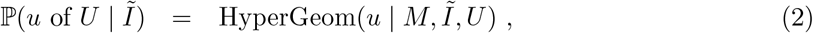

and this allows direct, accurate statistical inference on *Ĩ*, the proportion of the population that would return a positive PCR test.

#### Focusing prevalence on the infectious subpopulation

PCR tests are sensitive, and can detect the presence of SARS-CoV-2 both days before and weeks after an individual is infectious. It is usually desirable for prevalence to represent the proportion of a population that is infectious. We can obtain a likelihood for the number of infectious individuals *I* as follows:

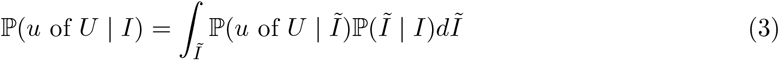

where *I* and *Ĩ* are the number of infectious and PCR-positive individuals respectively.

The conditional distribution ℙ(*Ĩ* | *I*) can be specified on the basis of external knowledge of the average length of time spent PCR-positive vs infectious. Our approach to estimating this quantity imports information on the timing of COVID-19 transmission [33] and the interval of PCR positivity in SARS-CoV-2 infected individuals [34]. More precisely, we specify the infectious time interval for an average infected individual in the population to span the interval 1 to 11 days post infection (the empirical range of generation time from Figure 1A of [33]). We then calculate the posterior probability of a positive PCR occurring 1 to 11 days post-infection (Figure 1A of [34]). We incorporate the effects of changing incidence in the calculations; this is important because, for example, if incidence is rising steeply, the majority of people who would test PCR positive in the population are those that are relatively recently infected. Full details can be found in Supplementary Information–*PCR positive to infectious mapping – method details*

#### Targeted surveillance data, *n* of *N*

In contrast to the randomised surveillance likelihood at (2), the targeted likelihood can be expressed in terms of the observation of *n* of *N* positive targeted (e.g. Pillar 1+2) tests as follows:

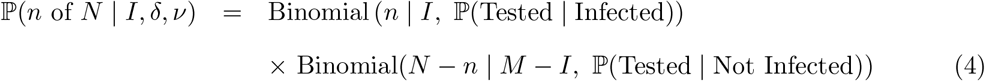

where ℙ(Tested | Infected) and ℙ(Tested | Not Infected) are the probabilities of an infected (respectively non-infected) individual being tested on date *t*.

#### Bias parameters, *δ* and *ν*

We introduce the following parameters:

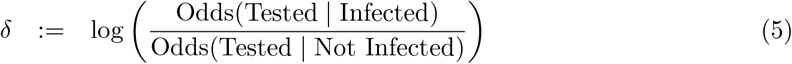

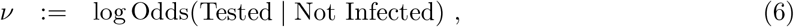

leading to the targeted swab testing likelihood being represented as

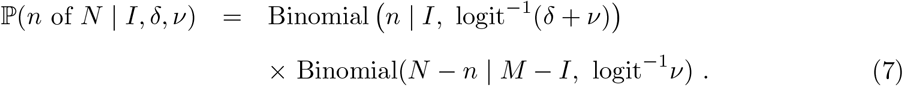

The unknown parameter requiring special care to infer is *δ*, i.e. the log odds ratio of being tested in the infected versus the non-infected subpopulations. The other parameter, *ν*, is directly estimable from the targeted data: 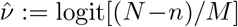 is a precise estimator with little bias when prevalence is low.

#### Test sensitivity and specificity

The likelihood at (7) assumes a perfect antigen test. If the test procedure has false-positive rate *α*, and false-negative rate *β*, the targeted likelihood is instead

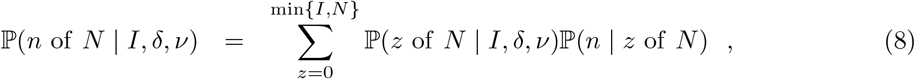

where *z* denotes the unknown number of truly infected individuals that were tested. The first term in the sum at (8) is obtained by substituting *z* in (7), while the second term is

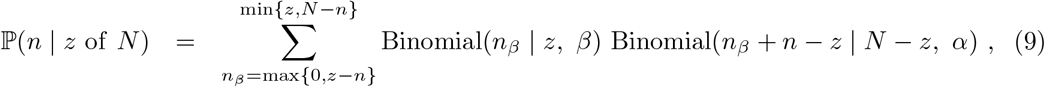

with *n*_*β*_ denoting the number of false-negative test results. An analogous adjustment can be made to the randomised surveillance likelihood at (2).

### Cross-sectional inference on local prevalence

We leverage spatially coarse-scale randomised surveillance data to specify an EB prior on bias parameters *p*(*δ*) at coarse-scale (PHE region), and thereby infer prevalence accurately from targeted data at fine scale (LTLA *j* within PHE region *J*_*j*_). We explicitly use the superscripts LTLA (*j*) in PHE region (*J*_*j*_) in step 4 below where notation from both coarse and fine scale appear together. All quantities in steps 1-3 are implicitly superscripted (*J*_*j*_) but these are suppressed for notational clarity. For computational efficiency we handle prevalence in a reduced-dimension space of bins as described in Supplementary Information (SI) section *Interval-based prevalence inference – set-up and assumptions*. The method in detail is as follows:

1. **Infer prevalence from unbiased testing data**. At a coarse geographic level (PHE region *J*_*j*_), estimate prevalence from randomised surveillance data *u*_*t*_ of *U*_*t*_. Represent the posterior at time *t* in mass function

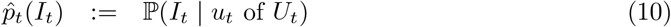

where 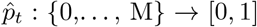 need only be available at a subset *t* ∈ 𝒯 ⊆ {1, …, *T*} of time points.
2. **Learn** *δ*_*t*_ **from accurate prevalence**. At a coarse geographic level, for each *t* ∈ 𝒯, we estimate bias parameter *δ*_*t*_ by coupling biased data *n*_*t*_ of *N*_*t*_ with accurate prevalence information 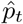. With *ν*_*t*_ fixed at 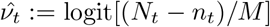

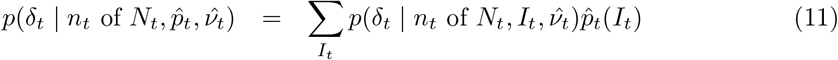

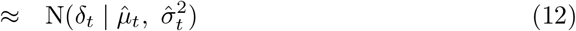

where a moment-matched Gaussian approximation is performed at (12). The posterior density in the sum at (11), *p*(*δ*_*t*_ | *n*_*t*_ of *N*_*t*_, *I*_*t*_, 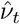) is conjugate under a Beta(*a,b*) prior on logit^−1^(*ν*_*t*_ + *δ*_*t*_) ≡ ℙ(Tested | Infected), and so can be evaluated as

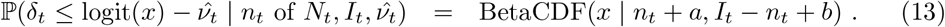
3. **Specify smooth EB prior on** *δ*_1:*T*_. A smooth prior on *δ*_1:*T*_ is specified as follows

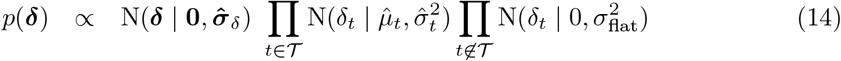

where N(***δ*** | **0, Σ**_*δ*_) imparts a user-specified degree of longitudinal smoothness, thereby sharing information on *δ* across time points. Ignorance of *δ*_*t*_, in the absence of random surveil-lance data, is encapsulated in a Gaussian with large variance 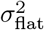. A standard choice for N(***δ*** | **0, Σ**_*δ*_) corresponds to a stationary autoregressive, AR(1), process of the form

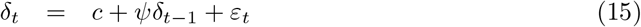

with a diffuse Gaussian prior 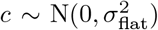 and with smoothing tuned by 0 *< ψ <* 1 and white noise variance 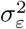. The normalised form of the prior at (14) is

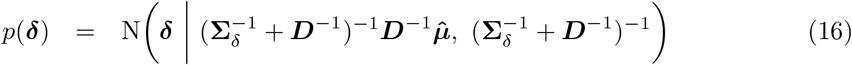

with (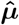, diagonal matrix ***D***_*T ×T*_) having elements 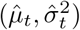 for *t* ∈ 𝒯 and 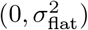 for *t* ∉ 𝒯.
4. **Infer cross-sectional local prevalence from biased testing data**. At a fine-scale geographic level (LTLA *j* in PHE region *J*_*j*_), having observed 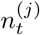 of 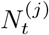 positive test results (a subset of the 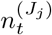 of 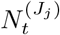 observed at the coarse-scale level above), calculate the posterior for 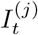 separately at each time point *t*:

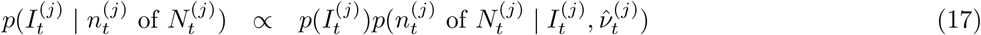

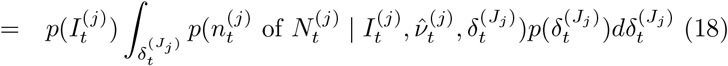

where 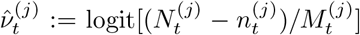, the likelihood in the integral at (18) is available at (7), and the prior 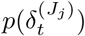 is time-point *t*’s marginal Gaussian from (16).

### Debiasing lateral flow device (LFD) tests with PCR surveillance (or vice versa)

The methods can be adapted straightforwardly to the situation in which the randomised surveillance study uses a different assay to the targeted testing. For a concrete example we could use REACT PCR prevalence posterior 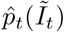 from (10) to debias Pillar 1+2 LFD test data *n*_*t*_ of *N*_*t*_. Equation (11) can be adjusted to estimate ascertainment bias *δ* pertaining to LFD data as follows:

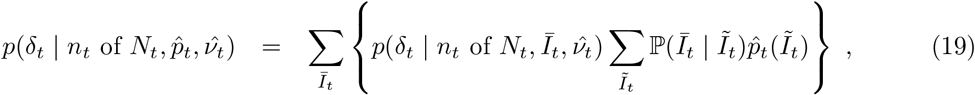

where *Ī*_*t*_ and *Ĩ*_*t*_ are the unobserved LFD- and PCR-positive prevalence respectively, and the conditional distribution ℙ(*Ī*_*t*_ | *Ĩ*_*t*_) can be estimated on the basis of external knowledge of the average length of time spent PCR-positive vs LFD-positive, analogously to as described in Methods– *Focusing prevalence on the infectious subpopulation*. The remaining computations, from (12) onwards, are unchanged, with the outputted fine-scale marginal likelihood 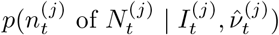 at (17) to be interpreted as targeting the local LFD-positive prevalence 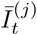.

### Full Bayesian inference under a stochastic SIR epidemic model

The cross-sectional analysis described in *Cross-sectional inference on local prevalence* generates the *δ*-marginalised likelihood, 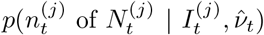 at (17), at each time point for which targeted data are available. These likelihoods can be used as input for longitudinal models to obtain better prevalence estimates and to infer epidemiological parameters such as *ℛ*_*t*_.

We illustrate this via a Bayesian implementation of a stochastic epidemic model whereby individuals become immune through population vaccination and/or exposure to COVID-19 (Figure 11). We incorporate known population vaccination counts into a standard discrete time Markov chain (DTMC) SIR model ([35], Chapter 3). Details of the transition probability calculations are given in SI section *SIR model details*, and assumptions in Supplementary Information–*SIR model – discussion, assumptions and caveats*.

**Figure 11:**
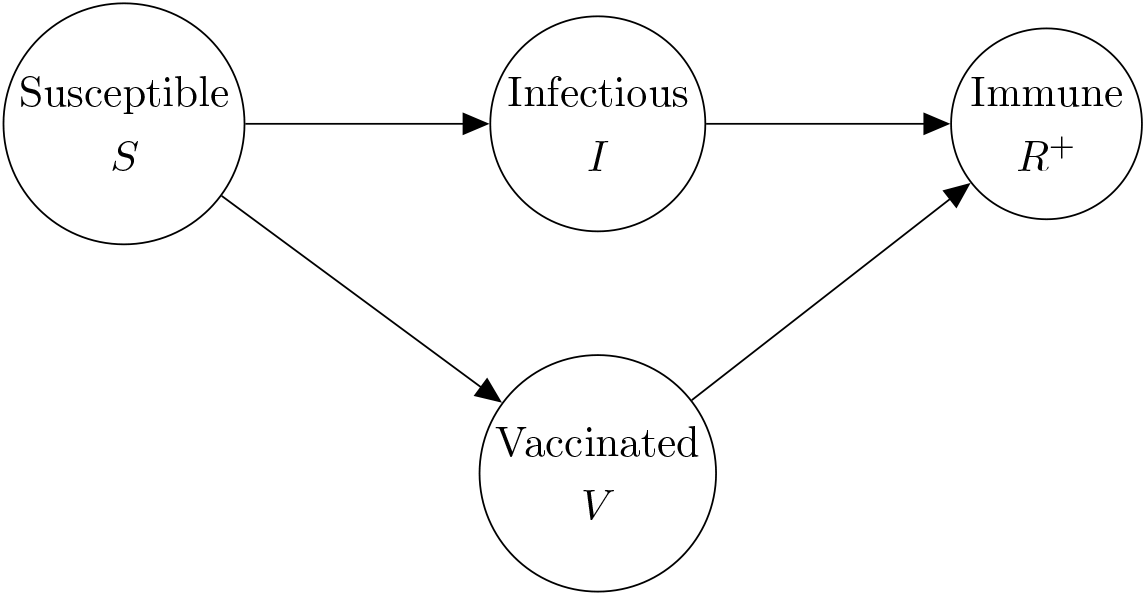
SIR/V epidemic model compartmental diagram.

### Priors on *ℛ, I, R*^+^

We place priors on ***I, R***^**+**^ measured as a proportion of the population; this proportion then gets mapped to prevalence intervals on subpopulation counts as described in *Interval-based prevalence inference – set-up and assumptions*. Specifically, we use truncated, discretized Gaussian distributions on the proportion of the population immune and infectious. For example, on number of infectious individuals *I*_*t*_ at each timepoint *t*, we specify the prior (suitably normalized over its support)

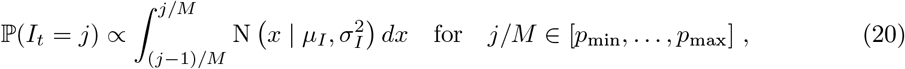

with an example weakly informative hyperparameter setting being *µ*_*I*_ = 0.5%, *σ*_*I*_ = 1%, *p*_min_ = 0%, *p*_max_ = 4%. To ensure meaningful inference on 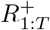, we place an informative prior that reflects the state of knowledge of the immune population size; we do this using an informative truncated Gaussian prior on 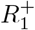, and non-informative priors on 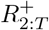. We place a noninformative uniform prior on each *ℛ*_*t*_, e.g. a Uniform(0.5, 2.5).

#### MCMC sampling implementation

We perform inference under the model represented in the DAG at Figure 11. The likelihood is marginalised with respect to *δ*, and we use Markov chain Monte Carlo (MCMC) to draw samples from the posterior

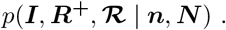

We sample ***ℛ*** and (***I, R***^**+**^) using separate Gibbs updates. For sampling (***I, R***^**+**^) we represent the joint full conditional as

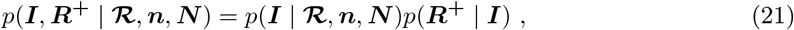

sampling ***I***^new^ from *p*(***I*** | ***ℛ, n, N***), and then 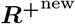 from *p*(***R***^**+**^ | ***I***^new^).

##### Sampling from *p*(*I* | *ℛ, n, N*)

The sampling distribution on prevalence can be expressed:

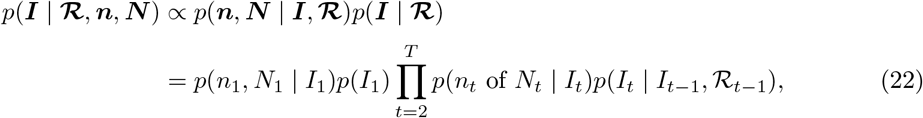

which is an HMM with emission probabilities taken from the *δ*-marginalised likelihood at (18), and transition probabilities taken from (37).

##### Sampling from *p*(*R*^+^ | *I*)

We can express the full conditional for 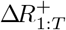 as

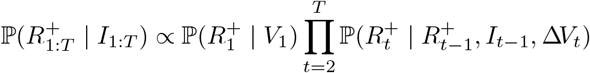

and sample the 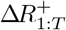 sequentially, with 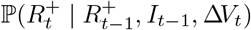 available at (39).

##### Sampling from *p*(*ℛ* | *I*)

The prior joint distribution of *ℛ*_1:*T*_ is modelled using a random walk:

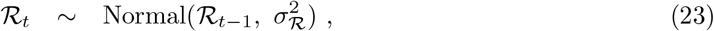

where 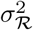 is a user-specified smoothness parameter.

The update involves sampling from

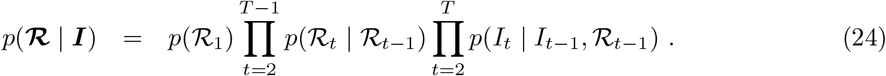

We discretize the space of *ℛ*_*t*_ into an evenly spaced grid and sample from the HMM defined at (24) [36]. The transition probabilities are given by (23) (suitably normalised over the discrete *ℛ*_*t*_ space) and the emission probabilities given by (37).

### Data

With the exception of the variant of concern 202012/01 analysis, all data underlying the results presented here are publicly available. Randomised surveillance data comes from the REACT study [7].^7^ From REACT, we create weekly test counts at the spatially coarse-scale level (PHE region) and, for validation purposes but not model fitting, use round-aggregated counts at the fine-scale level (LTLA), for round 7 (13th Nov - 3rd Dec 2021) and round 8 (6th-22nd Jan 2021). The combined weekly Pillar 1 and Pillar 2 data are publicly available for download.^8^

### Analysis scripts

The R scripts used to generate the results in this manuscript are available at https://github.com/alan-turing-institute/jbc-turing-rss-testdebiasing.

## Discussion

We have introduced and applied an integrative causal model allowing accurate inference from community testing data. The flexible probabilistic framework allows simultaneous and coherent incorporation of a number of important features, including:

- Adjustment for ascertainment bias caused by preferential testing based on symptom status, or on other confounders.
- Allowing for heterogeneous testing capacity by modelling the total number of tests conducted locally.
- Incorporation of multiple different SARS-CoV-2 testing assays, such a LFD and PCR, including adjustment for particular sensitivity/specificity.
- Inference on the number of infectious individuals, when PCR tests pick up individuals at non-infectious stages.
- The model outputs week-specific debiased prevalence with uncertainty (via a marginal likelihood), that can be incorporated modularly into more complex models.
- An SIR epidemic model implementation allowing estimation of *ℛ*_*t*_ and adjustment for vaccination in the immune population

Because of the extensive Pillar 1+2 testing effort, there is a large amount of information contained in these targeted data at LTLA level, even for a single weekly timepoint, as shown by the narrow width of CIs in Figure 4. However, due to the strength of this information, the targeted data also have the potential to introduce bias into more complex models if they are incorporated as observed nodes without an appropriate ascertainment correction. Equipped with a well specified prior on *δ*, however, precise and accurate estimation at even finer scales may be feasible. Given the high volume of data, despite focusing on high resolution geographical units such as LTLAs, estimates do not seem to be affected by the classical issue of small area estimation, that is to say sample sizes too small to carry on inference without borrowing strength from neighbouring units. If interest is focused on ultra fine-scale geography and/or when prevalence is much lower, then spatial borrowing would be beneficial, with such additional smoothing subject to a variance-bias trade-off.

In addition to estimating at finer spatial scales, it may be desirable to estimate local prevalence from targeted data stratified by factors such as age and ethnicity. This would involve modelling the relevant confounders in Figure 1, rather than marginalising them out. One approach to doing this within the existing framework would be to perform a stratified analysis (e.g. performing the whole analysis using weekly PHE region and Pillar 1+2 Data stratified by age bands). A more sophisticated approach could model *δ*_*t*_ semi-parametrically with some spatiotemporal smoothness assumptions on the effect of age and other confounders. Of course, any approach would require appropriate metadata on any confounders that affect randomised surveillance and/or targeted sampling.

For cross-sectional prevalence estimation, a key dependency is the availability of a regular, up-to-date stream of randomised surveillance data at some level of spatial resolution. Here we deployed REACT data at the coarse PHE-region scale. The UK has led the way internationally in having regular national surveillance randomised surveys like REACT and ONS. This modelling work shows the importance of having both targeted testing and also a rolling randomised surveillance survey to be able to better track the epidemic. The methods are transferable beyond the UK wherever randomized testing data are being gathered. can be applied in other This could be built in an integrated way from the start as preparedness for pandemics, in particular for diseases where asymptomatic transmission plays an important role.

## Data Availability

With the exception of the variant of concern 202012/01 analysis, all data underlying the results presented here are publicly available. Randomised surveillance data comes from the REACT study. From REACT, we create weekly test counts at the spatially coarse-scale level (PHE region) and, for validation purposes but not model fitting, use round-aggregated counts at the fine-scale level (LTLA), for round 7 (13th Nov - 3rd Dec 2021) and round 8 (6th-22nd Jan 2021). The combined weekly Pillar 1 and Pillar 2 data are publicly available for download.

https://github.com/mrc-ide/reactidd/tree/master/inst/extdata

https://www.gov.uk/government/publications/nhs-test-and-trace-england-statistics-14-january-to-20-january-2021

## Supplementary Information

### Model parameters

A full list of model parameters, along with either their prior distribution or the value at which they were fixed, can be found in Table S1.

**Table 1:** Model parameters with specified prior distributions or fixed values

| Parameter | Prior / Fixed value |
| --- | --- |
| Ascertainment bias, $\delta_{1:T}$ | Empirical Bayes prior (see Eq. (16)):<br>- AR(1) coefficient, $\psi = 0.99$<br>- Standard deviation, $\sigma_\epsilon = 1$<br>- Intercept, $c \sim \mathcal{N}(0, \sigma_{flat}^2)$ with $\sigma_{flat} = 10$ |
| PCR false-positive rate, $\alpha$ | Fixed, $\alpha = 0.001$, taken from [37, 38] |
| PCR false-negative rate, $\beta$, | Fixed, $\beta = 0.05$, taken from [39] |
| Expected time to recovery, $1/\gamma$ | $T_{\text{recovery}} \sim \text{Exponential}(\gamma)$, with $\gamma = 1$ week |
| Effective reproduction number, $\mathcal{R}_t$ | Random walk: $\mathcal{R}_t \sim \mathcal{N}(\mathcal{R}_{t-1}, \sigma_{\mathcal{R}}^2)$, with $\sigma_{\mathcal{R}}^2 = 0.2$ |
| Proportion immune at $t = 0$, $R_0^+/M$ | Truncated Gaussian (see Eq. (20), and reference [40])<br>- Mean, $\mu_R = 0.06$<br>- Standard deviation, $\sigma_R = 0.01$<br>- Minimum proportion, $p_{\min} = 0$<br>- Maximum proportion, $p_{\max} = 0.1$ |
| Proportion infectious at each $t$, $I_t/M$ | Truncated Gaussian (see Eq. (20)):<br>- Mean, $\mu_I = 0.005$<br>- Standard deviation, $\sigma_I = 0.01$<br>- Minimum proportion, $p_{\min} = 0$<br>- Maximum proportion, $p_{\max} = 0.04$ |

## Discussion of methodological assumptions and caveats

### Interval-based prevalence inference – set-up and assumptions

The full prevalence state space comprises all potential numbers of infectious individuals in the population, i.e. *I* ∈ {0, …, *M*}. For computational tractability we define *B* ≪ *M* bins:^9^

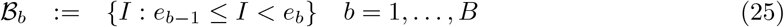

having midpoints:

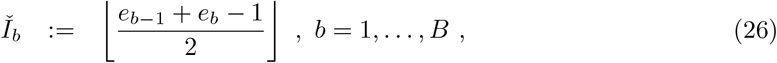

and make three assumptions to allow computationally efficient inference on the *B*-dimensional space of bins, denoting these assumptions Interval-1:3 as follows:

Interval-1 The testing data likelihood, conditional on prevalence bin, is evaluated at the bin mid-point:

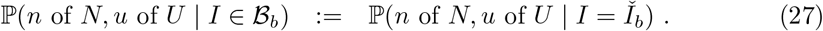

Interval-2 Prevalence *I* is uniformly distributed within each bin:

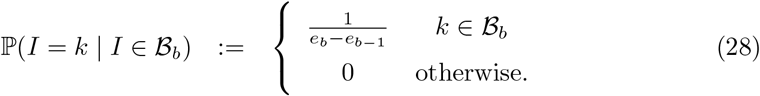

Interval-3 The distribution of new infections, conditional on prevalence bin, is evaluated at the bin midpoint (with the same assumption applying to new recoveries):

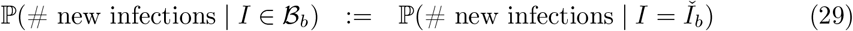

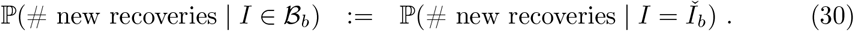

### Ascertainment bias model – assumptions and caveats

Debias-1 Spatial homogeneity of *δ* across LTLAs within a PHE region. The fact that we see relatively low variation in *δ* at each time point across PHE regions in Figure 3, particularly after October 2020, is consistent with a finer-scale spatial homogeneity assumption being reasonable.

Debias-2 We handle prevalence in a reduced-dimension space of bins as described in SI section *Interval-based prevalence inference – set-up and assumptions*

Debias-3 (In)stability of ascertainment mechanism. It is clear from Figure 3 that the ascertainment effects captured by *δ* can change rapidly and without obvious cause over time. Contemporaneous randomised surveillance data, such as REACT or ONS CIS, allow estimation of *δ*. However, when predicting prevalence forward in time beyond availability of randomised surveillance data, we are making the implicit assumption that the ascertainment bias remains stable forwards in time, and such results should therefore be interpreted with caution.

### PCR+ to infectious mapping – assumptions and caveats

For full details please see Supplementary Information—*PCR positive to infectious mapping – method details*.

Infectious-1 Pillar 1+2 positive test counts, across a four-week period, are used as an approximation to the true *relative* incidence over that time interval at coarse-scale level (e.g. PHE region).

Infectious-2 The probability (with credible intervals) of testing PCR positive when swabbed *d* days post infection is taken from Figure 1A of Hellewell et al. [34].

Infectious-3 The infectious interval for an average individual is defined to span days 1 to 11 post infection, based on Figure 1A of Ferretti et al. [33].

### SIR model – discussion, assumptions and caveats

The illustrative epidemic model we implement here has one of the simplest SIR compartmental structures available, as summarised in Supplementary Information–*SIR model – discussion, assumptions and caveats* and particularly Assumption SIR-2. Other teams have developed more realistic and sophisticated compartmental models of transmission, reflecting for example that individuals are not immediately infectious after being infected [22, 41, 42, 43]. Importantly, these are able to relate epidemiological disease dynamics to outcomes far downstream, such as hospitalisation and deaths. The fact that a large number and variety of models has been developed can be viewed as a strength, as demonstrated by efficacy of ensembles of multi-model forecasts to inform policy on future resource needs and population impacts [19]. One attractive feature of such model ensembles is that their forecasts may be relatively robust to changes in spatiotemporal and compartmental dynamics over the course of an epidemic. Notably, the de-biased prevalence likelihood outputted in Results–*Cross-sectional local prevalence from targeted testing data* is agnostic to the downstream epidemic model, and so there might be benefits to incorporating it into such multi-compartment epidemic models.

SIR-1 The population is homogeneous within an LTLA, with each individual equally likely to be infected

SIR-2 We assume individuals become instantly infectious and recover at a fixed rate *γ* = 7 days, i.e. with no spatiotemporal variation, and with recovery time distributed exponentially with mean 1*/γ*.

SIR-3 Any projections forward in time are made under the implicit assumption that there is no change in NPIs, such as tiering or lockdown status, affecting the LTLA.

SIR-4 We do not include age, ethnicity or deprivation indices in our model, and so epidemiological parameter estimates are to be interpreted as an average across these strata (with unknown weights).

SIR-5 We do not explicitly model transmission between regions or the demographic effects of births, deaths and migration – the SIR model is fitted to each LTLA separately. While it would be possible to account for transmission between LTLAs [44], this dramatically increases the number of parameters to be estimated and consequently the computational burden of the model. Given that the study period here is almost all in lockdown, the effect of transmission between LTLAs is relatively small. In non-lockdown periods, epidemic models allowing for inter-region transmission could be beneficial.

SIR-6 The number of new infections in the stochastic SIR model is modelled as a Poisson approximation, approximating the ‘true’ Binomial conditional distribution.

### Gaussian approximation for *δ*

We approximate the cross-sectional component of the EB prior for *δ* using a moment-matched Gaussian approximation (see (12)). Figure 13 illustrates the suitability of this approximation for PHE regions London and the North West across nine weeks.

**Figure 12:**
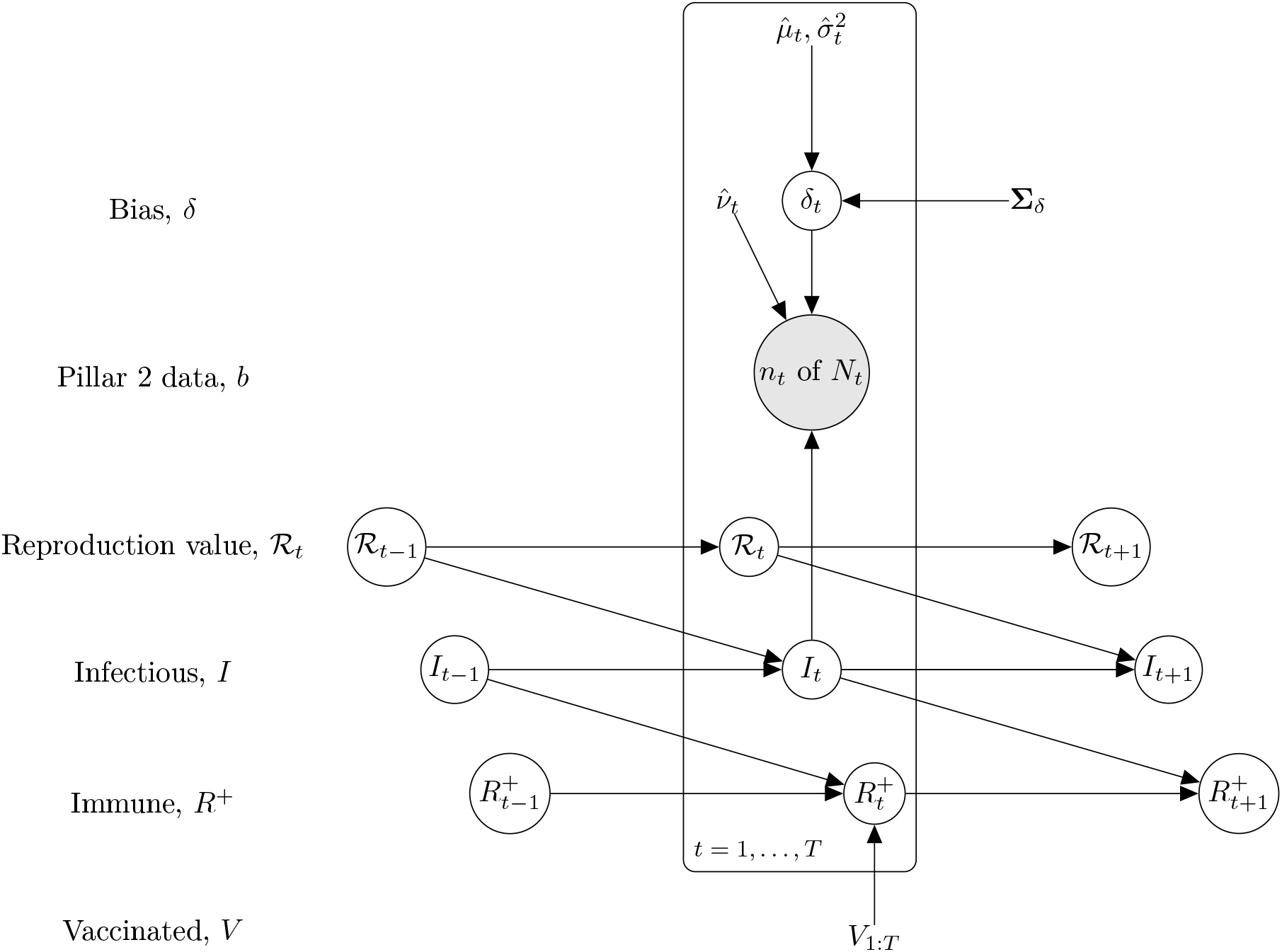
Longitudinal model DAG for SIR epidemic model at local level (e.g. LTLA). Directed paths characterise conditional probability distributions, in contrast to the paths showing transitions between model compartments in Figure 11. Inference is for a region, e.g. an LTLA, based only on targeted test data collected in this region, *n*_*t*_ of *N*_*t*_. A prior on *δ*_*t*_ parameterized 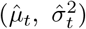 brings information on the Pillar 2 ascertainment bias learned from randomized surveillance testing data available for the PHE region in which the LTLA lies. The *T × T* covariance matrix **Σ**_*δ*_ imparts temporal smoothness on *δ*_1:*T*_. Effective reproduction numbers are denoted *ℛ*_1:*T*_, number of infectious individuals by *I*_1:*T*_, and the number of immune individuals by 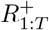.

**Figure 13:**
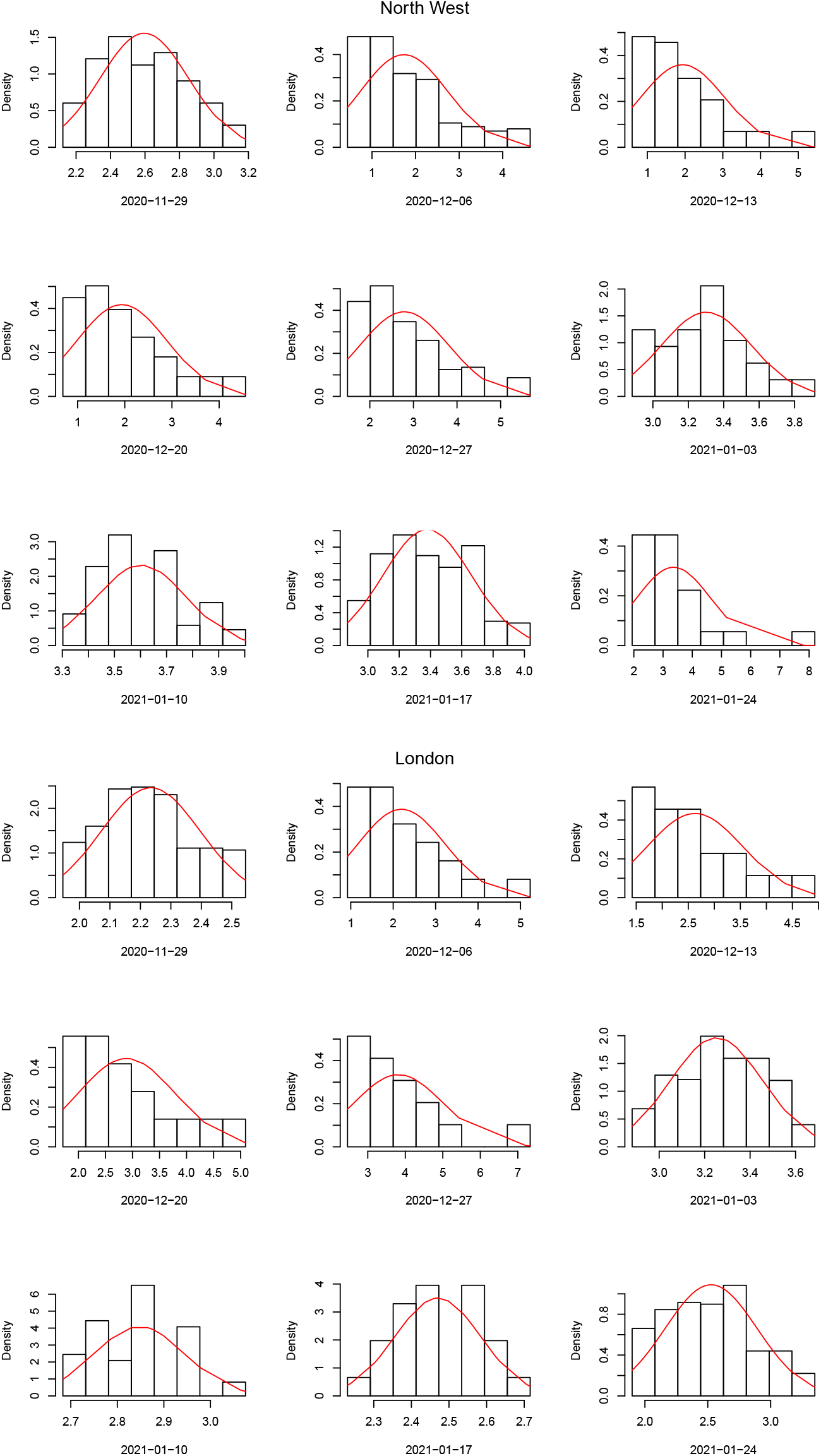
Comparison of moment-matched Gaussian EB prior (12) (red lines) with raw estimates (histograms) on *δ* for PHE regions North West (top) and London (bottom) from 29th November 2020 to 24th January 2021.

### SIR model details

We implement a DTMC SIR epidemic model based on the standard model as described in ([35], Chapter 3). As we choose Δ*t* to be a day/week, we allow multiple infections and recoveries in a time interval width Δ*t*; this requires derivation of Markov transition probabilities between all states (rather than just neighbouring ones), which we do below having established some notation.

### Notation

Parameters are subscripted by timepoint index *t* (indexing week for the analyses presented, with Δ*t* set to one week):

*I*_*t*_: number of infectious individuals

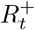: number of immune individuals (with infection- and/or vaccination-acquired immunity)

*V*_*t*_: total number of vaccinated individuals in region (i.e. with vaccine-acquired immunity)

*S*_*t*_: number of susceptible individuals 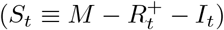

Δ*Q*_*t*_: number of *new* infections in interval (*t* − Δ*t, t*]

Δ*R*_*t*_: number of *new* recoveries in interval (*t* − Δ*t, t*]

Δ*V*_*t*_: number of vaccinations administered in interval (*t* − Δ*t, t*]

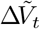: number of vaccinations administered to <u>*susceptible*</u> individuals in interval (*t* − Δ*t, t*]

*β*_*t*_: transmission rate, i.e. the number of effective contacts in interval (*t* − Δ*t, t*]

*γ*: recovery rate, with expected time to recovery 𝔼[*T*] = 1*/γ*

*γ*_*t*_: probability of recovery in interval (*t* − Δ*t, t*], i.e. *γ*_*t*_ := ℙ(*T* ≤ Δ*t*) where *T* ∼ Exp(*γ*)

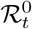: basic reproduction number, 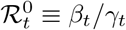

*ℛ*_*t*_: effective reproduction number, 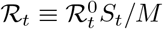

### Distribution of the number of new infections Δ*Q*_*t*_

Under the standard DTMC SIR model, the number of *new* infections, denoted here Δ*Q*_*t*_, occurring in the time interval Δ*t* up to time *t* has conditional distribution^10^

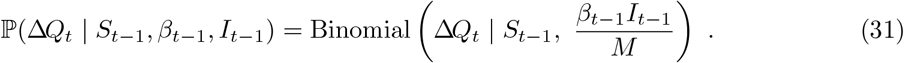

The probability in (31) can be parameterised by the effective reproduction number, *ℛ*_*t*_:

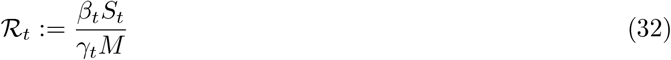

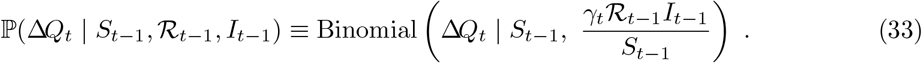

We approximate (33) with a Poisson distribution as follows [45]:^11^

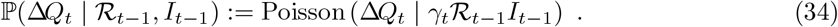

### Distribution of the number of new recoveries Δ*R*_*t*_

The number of *new* recoveries, denoted Δ*R*_*t*_, occurring in the time interval Δ*t* up to time *t* is distributed

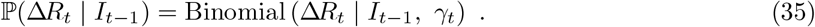

### Transition probabilities for the number of infectious individuals *I*_*t*_

The change in the number of infectious individuals at time *t*, Δ*I*_*t*_ can then be expressed as

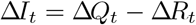

this and so the conditional distribution for Δ*I*_*t*_ follows from (34) and (35):

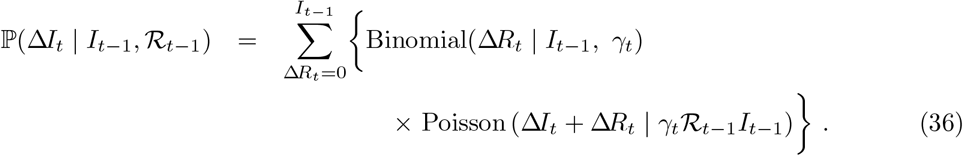

Interval-to-interval transition probabilities are evaluated as

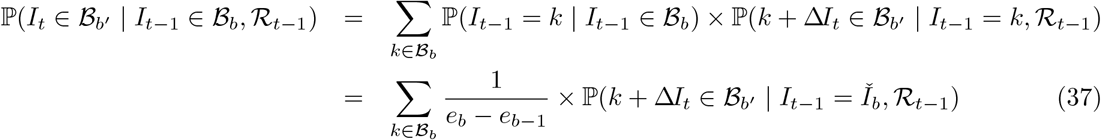

where the first term in the sum at (37) follows from Assumption 2 at (28), and the second term is conditional on prevalence at bin midpoint 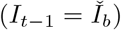 based on Assumption 3 at (29)-(30), and can be evaluated using (36).

### Transition probabilities for the number of immune individuals 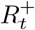

Denote by Δ*V*_*t*_ the number of vaccinations administered in interval (*t* − Δ*t, t*]. Only a subgroup of those individuals vaccinated at time *t* may have been susceptible at time *t* − Δ*t*; we denote the number in the subgroup by 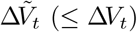, and evaluate its conditional distribution as follows:

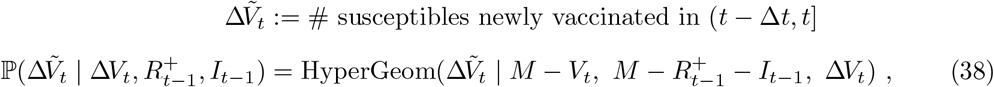

where *V*_*t*_ is the current number of vaccinated individuals in the population (with Δ*V*_*t*_ ≡ *V*_*t*_ − *V*_*t*−1_). The total number of immune, i.e. vaccinated and/or recovered, individuals at time *t* (denoted 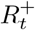) can then be represented by the recurrence

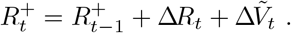

This leads to the Markov conditional distribution for 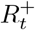 via convolution of (35) with (38)

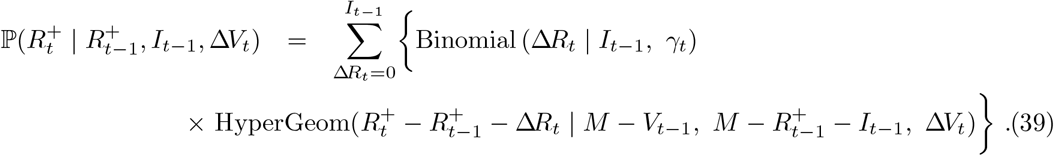

The above treatment of immunity assumes individuals are made permanently immune immediately through either vaccination or infection. It would be straightforward to relax the above formulation to allow for more sophisticated treatment of immunity, for example specifying (a) a delay in vaccine effects, (b) incomplete vaccine efficacy (e.g. in the case of novel variants), or (c) decaying immunity over time.

### Inference on the basic reproduction number

The basic reproduction number at time *t*, 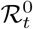 is related to the effective reproduction number *ℛ*_*t*_ by the following equation,

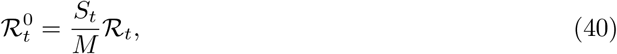

where *M* is the total number of individuals and *S*_*t*_ is the number of susceptible individuals at time *t*. Recall that 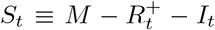 where 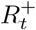 is the number of immune individuals and *I*_*t*_ is the number of infectious individuals, both of which are estimated by our DTMC SIR model. We can plug in these estimates into (40) to estimate 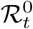 for a given LTLA. Figure 15 plots 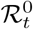 and *ℛ*_*t*_ for a selection of LTLAs.

**Figure 14:**
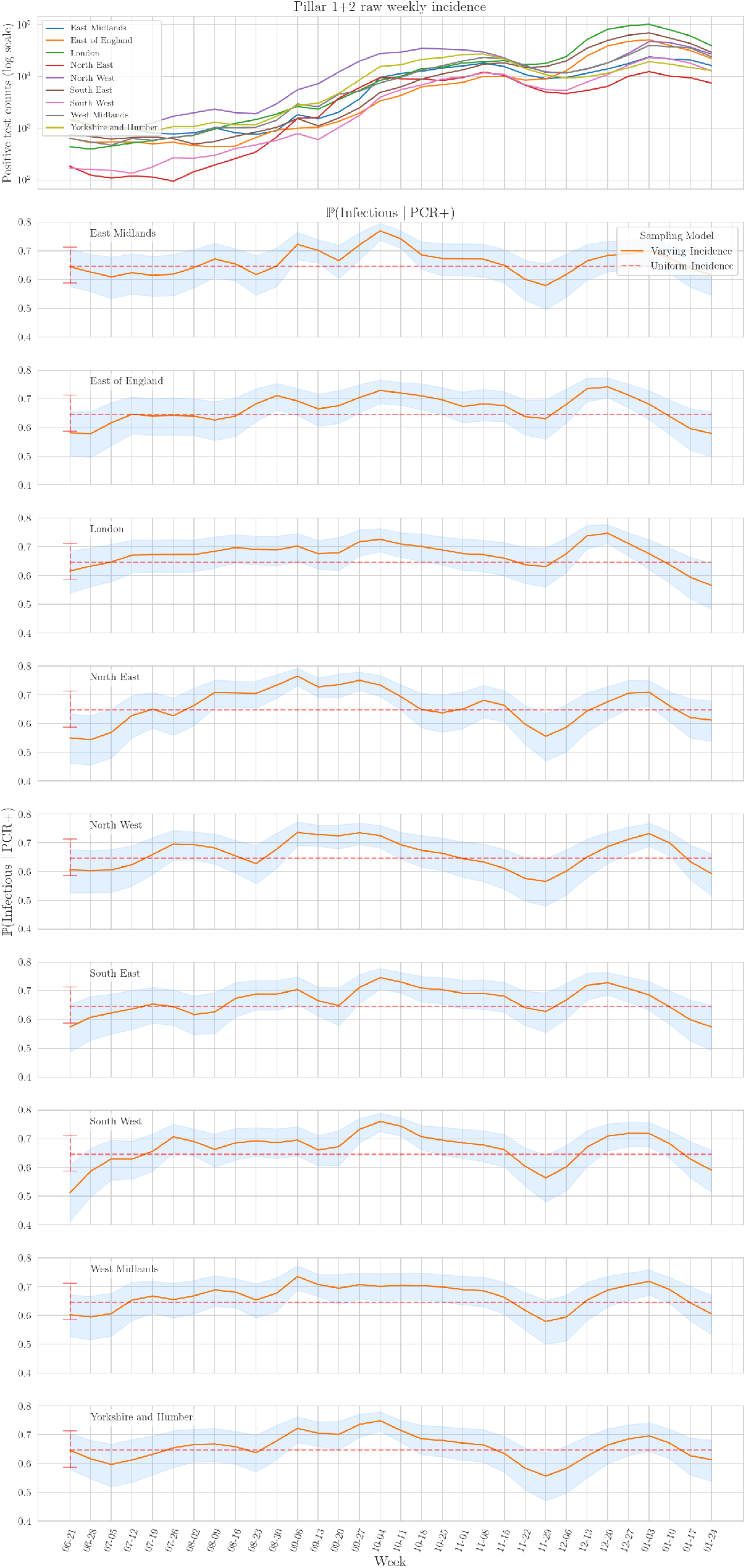
EB prior on ℙ(Infectious | PCR positive) by week and PHE region. The top panel shows raw weekly Pillar 1+2 incidence for the nine PHE regions; this is to provide intuition for the Varying incidence model in the panels below. The bottom nine panels display the prior we place on ℙ(Infectious_*t*_ | PCR+_*t*_), which is specific to week and region for the Varying incidence model, but is constant across weeks/regions for the Uniform incidence model (see legend in panel 2). Error bars (at left of panel for Uniform incidence; around curve for Varying incidence) represent 95% credible intervals.

**Figure 15:**
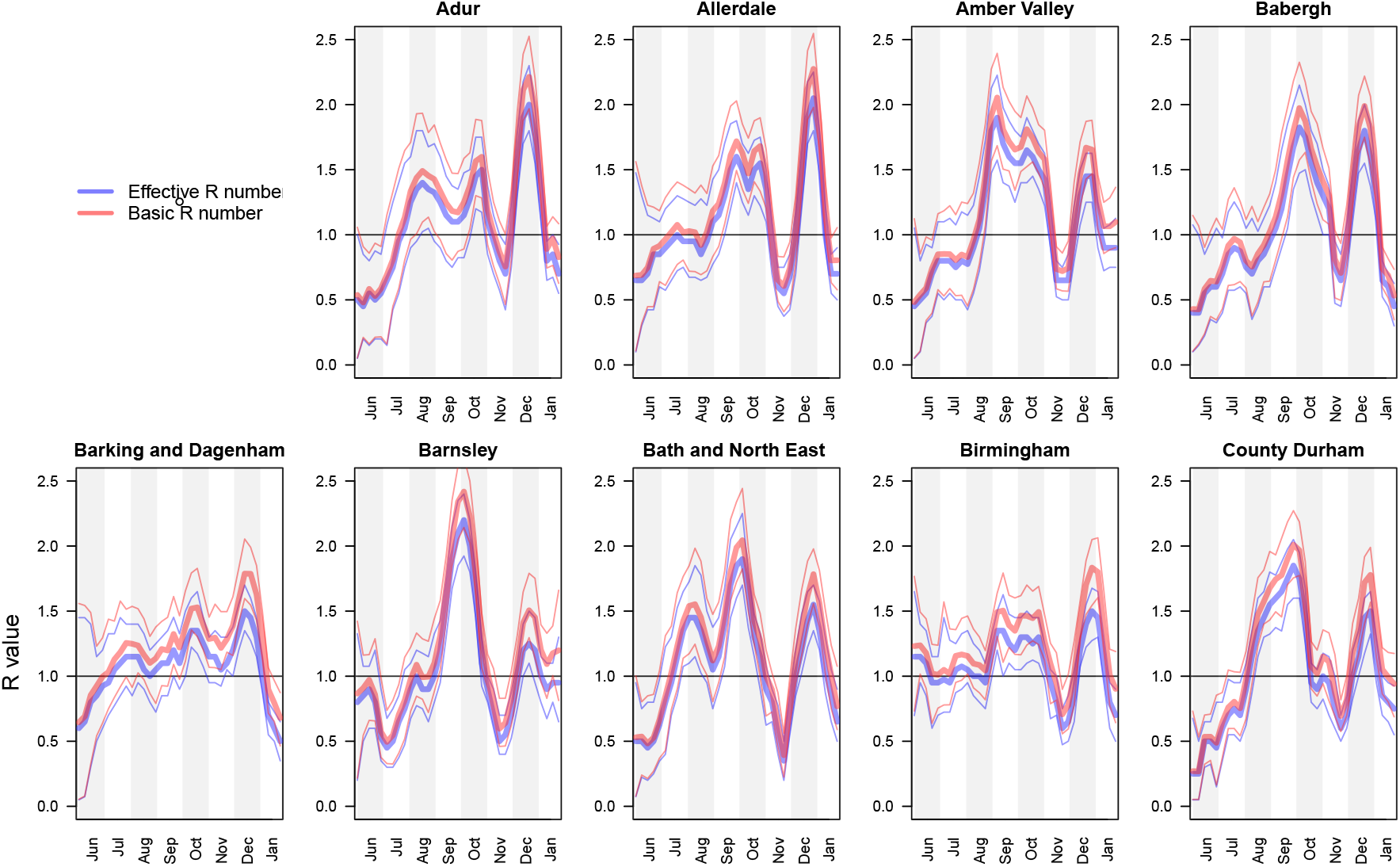
Comparison of 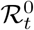 and *ℛ*_*t*_ estimates.

## PCR positive to infectious mapping – method details

Recall we require ℙ(*Ĩ* | *I*) in (3), which is the probability distribution on the number of PCR positive individuals *Ĩ* given the number of infectious individuals *I*. This can be expressed via Bayes’ theorem as

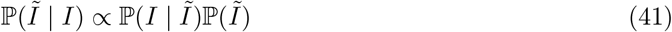

where the likelihood is binomial:

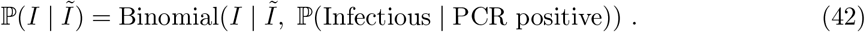

To target the ℙ(Infectious | PCR positive) success probability in (42), we introduce the following notation:

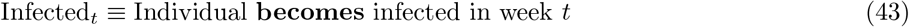

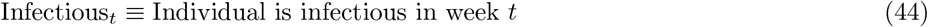

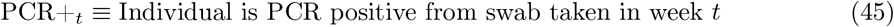

and proceed as follows:^12^

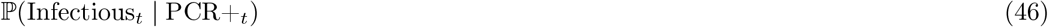

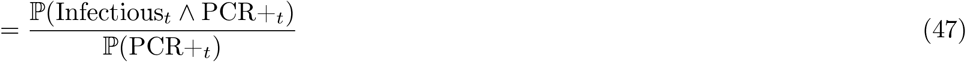

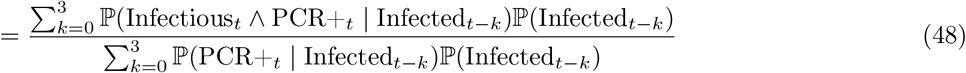

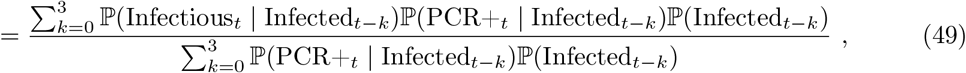

where, at (49), we assumed conditional independence between Infectious_*t*_ and PCR+_*t*_ conditional on Infected_*t*−*k*_. Also, at (48), we assumed that testing PCR positive implies that an individual was infected at most four weeks prior to being swabbed, which is consistent with Figure 1A of [34] (data input 2 below). We import three distinct data inputs to estimate the various terms in (49).

### Data input 1 – Infectious interval

Figure 1A of Ferretti et al. [33] shows the estimated probability density function of the serial interval for SARS-CoV-2 transmission – **we denote this density function** *f*_**Fer**_(*d*). Noting the support of this density to be approximately [1, 11], we specify that an average individual is infectious between days 1 to 11. Formally we define, independently for each individual in the population,

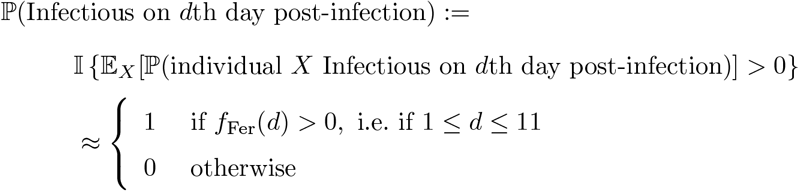

where *X* denotes an individual selected uniformly at random from the population. We can use this to estimate the ℙ(Infectious_*t*_ | Infected_*t*−*k*_) term appearing in the numerator of (49) as follows

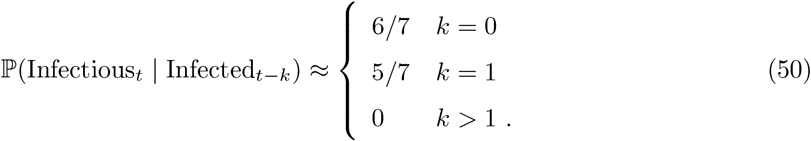

### Data input 2 – PCR positive interval

Figure 1A of Hellewell et al. [34] plots posterior probabilities (with credible intervals) of testing PCR positive when swabbed *d* days post infection. We denote this data input

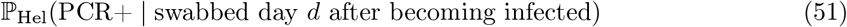

and use it to estimate the term ℙ(PCR+_*t*_ | Infected_*t*−*k*_) appearing twice in (49), evaluating the following estimator for each *k* = 0, …, 3:

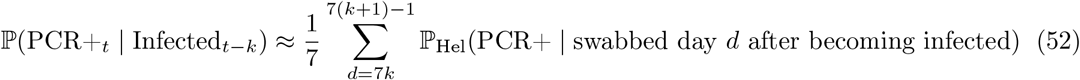

Hellewell et al. [34] helpfully provide reproducible scripts^13^ and we use these to extract the posterior distribution on ℙ_Hel_(PCR+ | swabbed day *d* after becoming infected) from their Figure 1A, whose uncertainty we propagate to estimator (52) and onwards to (49), yielding a distribution on ℙ(Infectious_*t*_ | PCR+_*t*_) which we take forward approximated by a moment-matched Beta distribution (at each week *t*) to be used as an EB conjugate prior on the success probability in (42).

### Data input 3 – Pillar 1+2 incidence

For the purposes of adjusting the PCR positive map to changing incidence, we use the raw regional weekly positive test counts *n*_0:*T*_, where we denote weeks by *t* = 0, …, *T*. We use this data input to estimate the term ℙ(Infected_*t*−*k*_) appearing twice in (49), evaluating the following estimator for each *k* = 0, …, 3:^14^

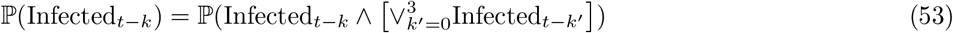

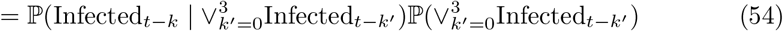

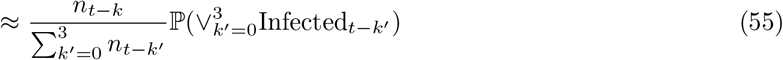

which can be directly substituted for ℙ(Infected_*t*−*k*_) in top and bottom of (49) with the second term on the right of (55) cancelling between numerator and denominator, and therefore not requiring evaluation. We note that we are using raw counts to model relative incidence over a relatively short period (four weeks), which is making the assumption that the bias is relatively stable over this timeframe (see Assumption Infectious-1 in SI–*PCR+ to infectious mapping – assumptions and caveats*).

### Sensitivity analyses

#### Prior hyperparameters for *δ*

The EB prior for *δ* depends on two hyperparameters: *σ*_*ϵ*_ controls the variance of the white noise associated with each individual time point, while *ψ* controls the degree of autocorrelation from one time point to the next. Figures 16 and 17 show the estimates for prevalence and *ℛ*_*t*_ respectively of infectious individuals using different values of these two hyperparameters. Note that in the main text, we present results using *σ*_*ϵ*_ = 1 and *ψ* = 0.99.

**Figure 16:**
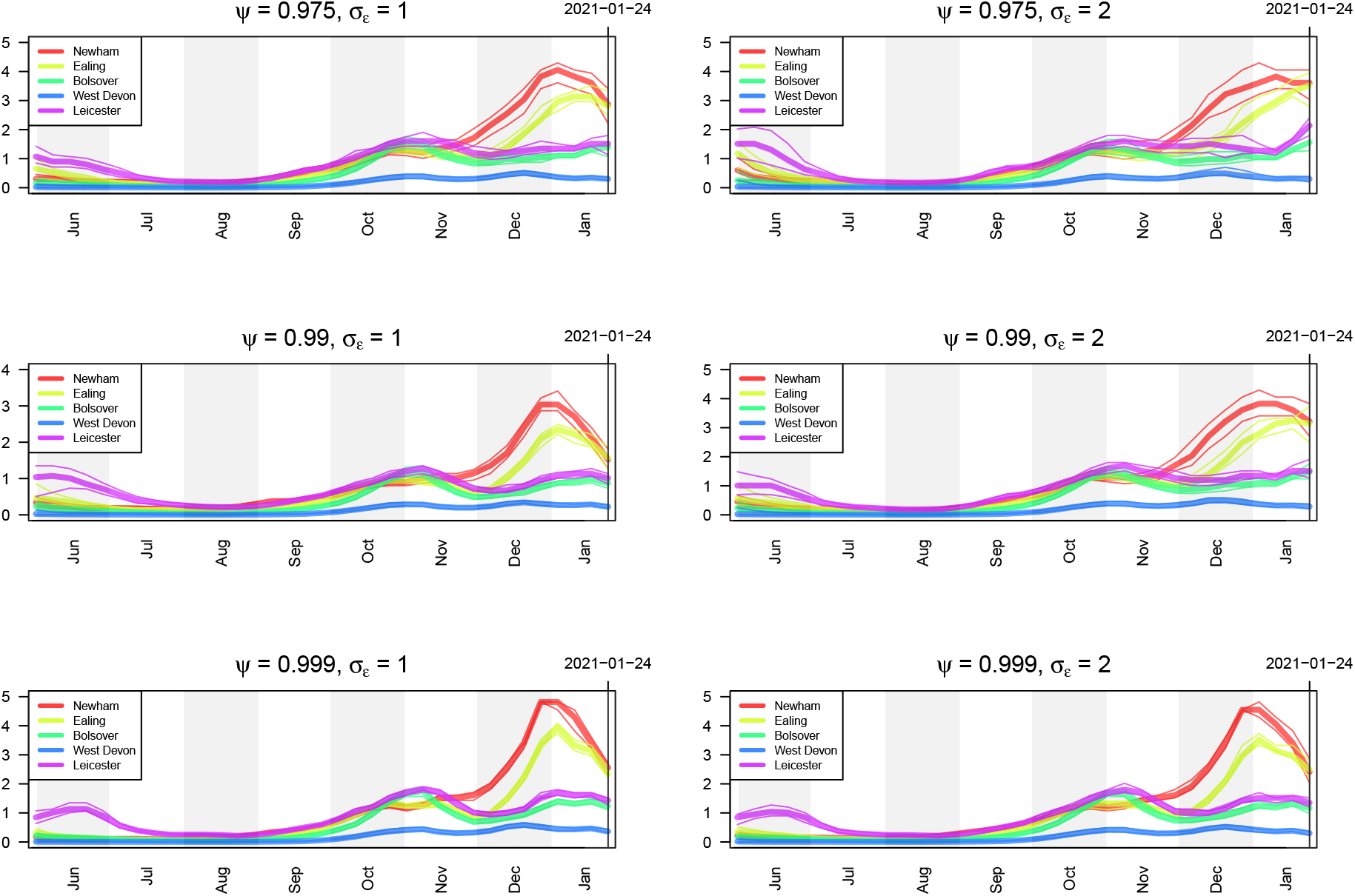
Estimates of prevalence of infectious individuals for five LTLAs using different values of the hyperparameters *σ*_*ϵ*_ and *ψ* controlling the smoothness of the bias parameter *δ*.

**Figure 17:**
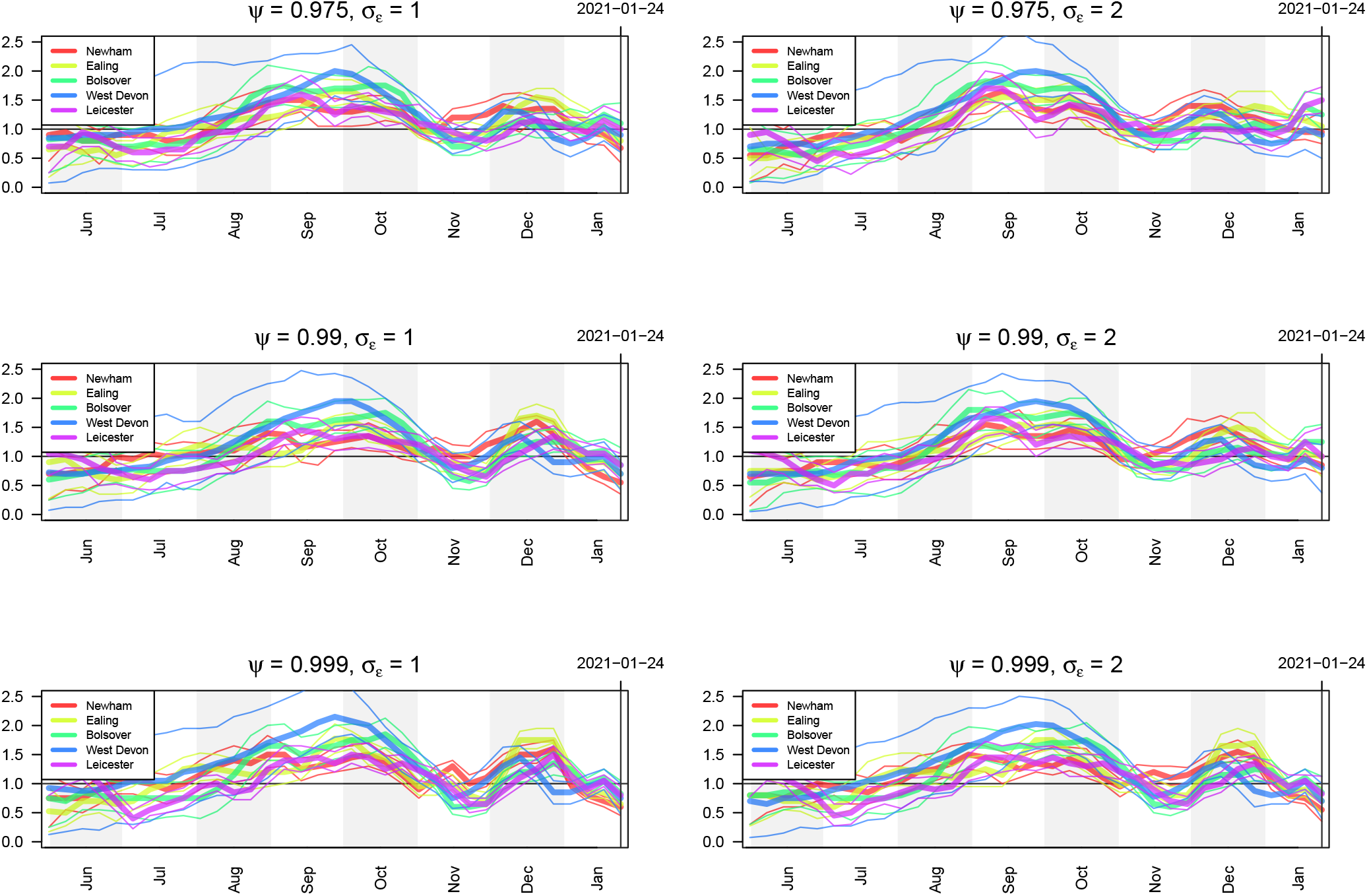
Estimates of prevalence of infectious individuals five LTLAs using different values of the false positive rate *α* and false negative rate *β* controlling the smoothness of the bias parameter *δ*.

#### Sensitivity and specificity of PCR tests

PCR tests are not perfect and are subject to both false positives and false negatives. In our analysis, we account for imperfect testing via the false positive rate, *α*, and the false negative rate, *β* (see (8)). Figures 18 and 19 show the estimates for prevalence and *ℛ*_*t*_ respectively of infectious individuals using different values of these two hyperparameters. Note that in the main text, we present results using *α* = 0.001 and *β* = 0.05.

**Figure 18:**
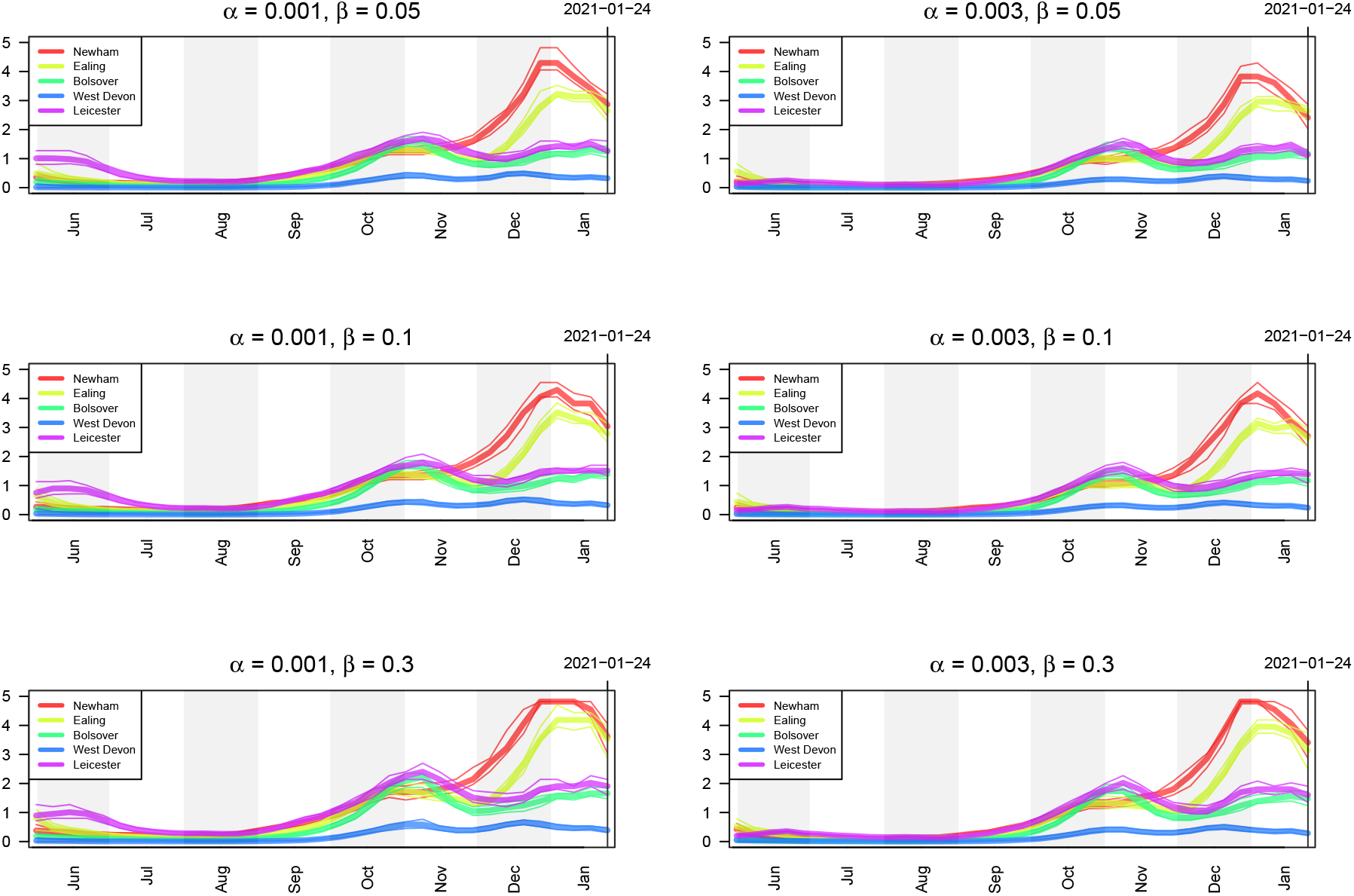
Estimates of *ℛ*_*t*_ of infectious individuals five LTLAs using different values of the false positive rate *α* and false negative rate *β* controlling the smoothness of the bias parameter *δ*.

**Figure 19:**
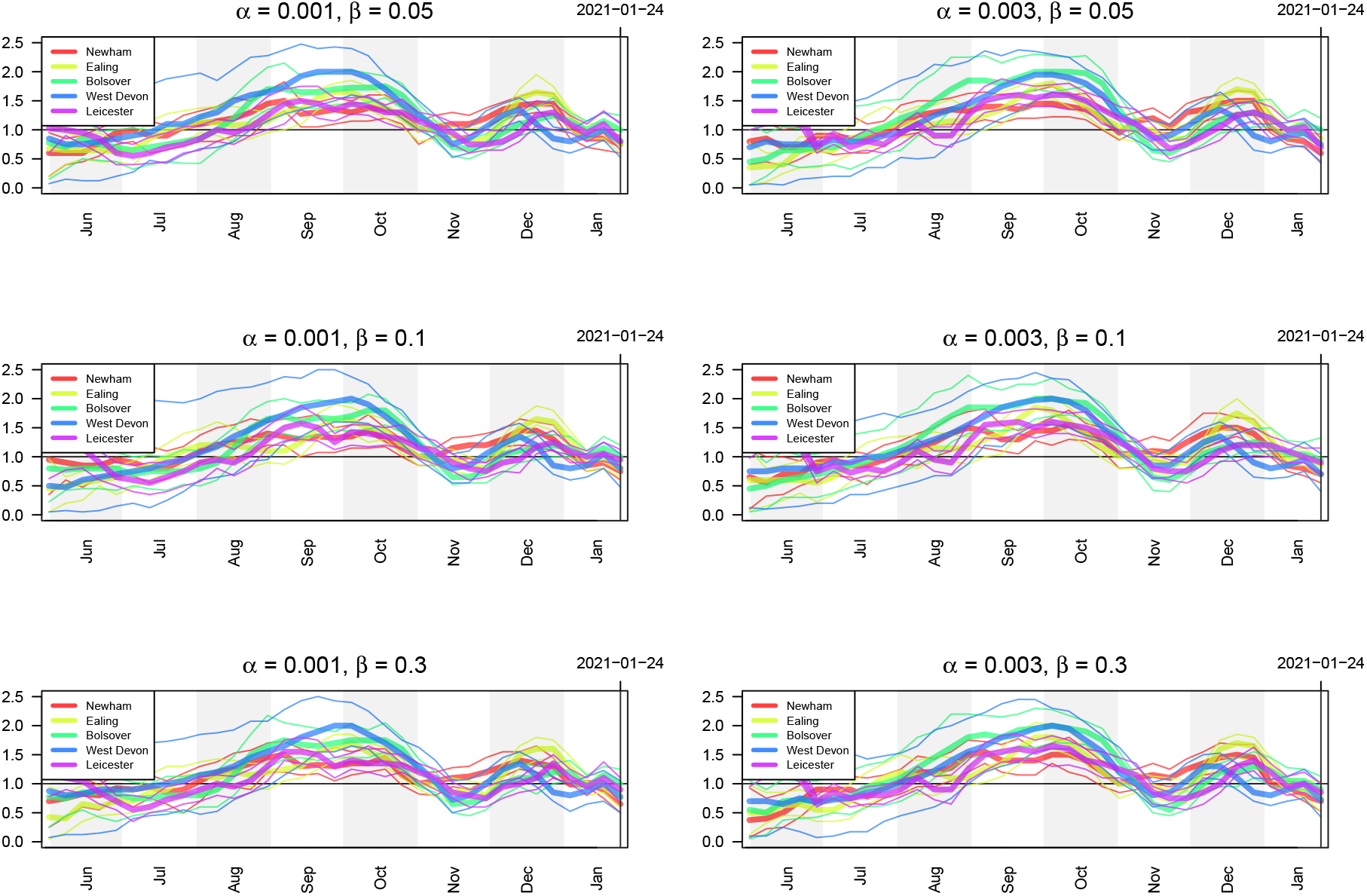
Estimates of prevalence of infectious individuals for five LTLAs using different values of the hyperparameters *σ*_*ϵ*_ and *ψ* controlling the smoothness of the bias parameter *δ*.

## Footnotes

1 Technical details of the methodology driving these websites is not yet available at the time of writing, but in both cases the peer-review process is underway.

2 https://localcovid.info/

3 https://rs-delve.github.io/

4 https://imperialcollegelondon.github.io/covid19local/#map

5 https://imperialcollegelondon.github.io/covid19local/#map

6 The imperial model uses daily cases data, weekly deaths data, as well as daily infections from the ONS CIS and REACT data sets

7 https://github.com/mrc-ide/reactidd/tree/master/inst/extdata

8 https://www.gov.uk/government/publications/nhs-test-and-trace-england-statistics-14-january-to-20-january-2021. Note that lateral flow test results are not included in the these weekly summaries.

9 Bins are equally sized on log scale, with interval edges are defined recursively as *e*_0_ = 0, *e*_*b*_ = ⌈*e*_*b*−1_(1 + *ε*_*B*_)⌉, and *ε*_*B*_ is a fixed constant giving *B* intervals.

10 Based on each of 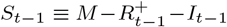 susceptibles at time *t*−1 being infected independently with probability ℙ(Susceptible infected | *β*_*t*−1_ effective contacts in (*t* − Δ*t, t*]) 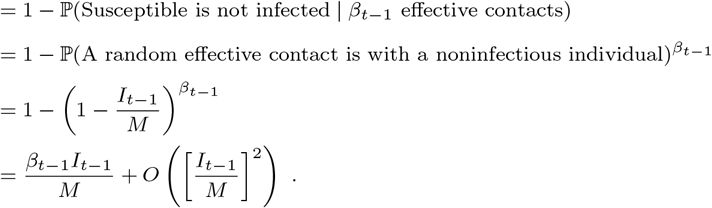

11 According to Rule 2 in [45], the Poisson approximation is reasonable when both of these inequalities hold: 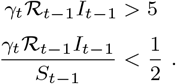 Of the two, the first is the least likely to obtain, but is still reasonable under most circumstances. For a simple example, if we set *γ*_*t*_ = 1 and *ℛ*_*t*−1_ = 1, the number of infectious individuals *I*_*t*−1_ *>* 5 is sufficient for the approximation to be reasonable.

12 We use ∧ to denote logical AND.

13 https://github.com/cmmid/pcr-profile

14 We use ∨ to denote logical OR.

